# Plasma pTau217 as a Prognostic, Risk-Stratification, and Monitoring Biomarker of Clinical Progression in Lewy Body Disease

**DOI:** 10.64898/2026.03.26.26349399

**Authors:** S. A. Lorkiewicz, C. Abdelnour, M. L. Bolen, A. M. Smith, M. Shahid-Besanti, D. Hemachandra, E. M. Müller-Oehring, N. Siddiqui, L. Montoliu-Gaya, B. Arslan, N. J. Ashton, E. N. Wilson, L. Tian, K. I. Andreasson, E. C. Mormino, V. W. Henderson, H. Zetterberg, K. L. Poston

## Abstract

**Background and Objectives:** While plasma phosphorylated tau-217 (pTau217) detects co-occurring Alzheimer’s disease (AD) neuropathologic change in Lewy body disease (LBD), relationships with long-term clinical outcomes remain unclear. Our aim was to evaluate plasma pTau217 as a prognostic, risk-stratification, and monitoring biomarker of cognitive and functional decline in LBD.

**Methods:** This prospective longitudinal study included Stanford Alzheimer’s Disease Research Center participants enrolled from 2015-2023 with plasma pTau217 data and clinical diagnoses of LBD spectrum, AD spectrum, or healthy control (HC). To evaluate prognostic and monitoring utility, respectively, linear mixed-effect models tested whether baseline level and longitudinal change (2-5 years) in plasma pTau217 were associated with longitudinal changes (2-8-years) in daily functioning (Clinical Dementia Rating-Sum of Boxes [CDR-SB]), global cognition (Montreal Cognitive Assessment [MoCA]), and 5 domain-specific cognitive indices. For risk-stratification, baseline plasma pTau217 status was defined using an amyloid PET-derived, LBD-specific cut-point to examine whether participants with abnormal levels demonstrated faster clinical progression in separate linear-mixed effects and survival models.

**Results:** A total of 501 participants (mean[SD] age = 71.1[8.5]; 51.5% female) were included across LBD (*n* = 131), AD (*n* = 133), and HC (*n* = 237) groups. In LBD, higher baseline plasma pTau217 was associated with faster CDR-SB increase (*β* = 0.30; 95% CI: [0.14, 0.45]; *p* < 0.001), MoCA decline (*β* = −0.35; 95% CI: [−0.57, −0.13]; *p* = 0.002), and cognitive index decline (all *p* ≤ 0.028). Participants with abnormal baseline pTau217 had a 0.85 points/year faster CDR-SB increase (95% CI: [0.56, 1.15]; *p* < 0.001), 0.87 points/year faster MoCA decline (95% CI: −1.38, −0.37; *p* = 0.001), faster decline on cognitive indices (all *p* ≤ 0.018), and a three-fold higher risk of diagnostically progressing to MCI or dementia (HR = 3.41; 95% CI: [1.60, 7.28]; *p* = 0.002) compared to participants with normal pTau217. Faster longitudinal pTau217 increase was associated with faster CDR-SB increase (*β* = 0.24; 95% CI: [0.10, 0.38]; *p* = 0.003).

**Discussion:** Plasma pTau217 is a promising prognostic, risk-stratification, and monitoring biomarker of clinical progression in LBD, underscoring its utility for clinical practice and trials in mixed pathology groups.

## Introduction

Lewy body disease (LBD) is the second leading cause of neurodegenerative dementia^1^ and encompasses a spectrum of disorders (e.g., Parkinson’s disease [PD], dementia with Lewy bodies) defined neuropathologically by intracellular aggregates of misfolded α-synuclein protein.^2^ Although abnormal α-synuclein deposition contributes to cognitive and motor dysfunction in LBD, cognitive decline is also associated with Alzheimer’s disease (AD) neuropathologic change (ADNC), defined as amyloid-β (Aβ) plaques and neurofibrillary tau tangles.^3,4^ ADNC is present in over 50% of individuals with LBD at autopsy,^5^ contributes to clinical heterogeneity and diagnostic uncertainty^6,7^, and is associated with worse clinical outcomes such as cognitive and functional decline, increased mortality, and reduced treatment response.^8–11^

Given the high prevalence and association with greater disease burden in LBD, in vivo identification of ADNC co-pathology has important implications for clinical practice and trial recruitment^1^. Indeed, blood-based biomarkers have emerged as clinically accessible and scalable tools for detecting ADNC mixed pathology in LBD.^12^ In particular, plasma phosphorylated tau at threonine 217 (pTau217), a validated diagnostic biomarker in AD,^13,14^ shows strong concordance with established biomarkers of ADNC co-pathology in LBD such as amyloid and tau positron emission tomography (PET) and cerebrospinal fluid (CSF) Aβ42/Aβ40 ratio.^15^ Recent studies further suggest plasma pTau217 can aid in diagnostic clarification in LBD,^16^ particularly when using LBD-specific thresholds to define abnormal levels^17^ and when considered alongside complementary plasma biomarkers.^18^

While PET, CSF, and genetic markers of co-occurring ADNC are associated with clinical progression in LBD,^4,19^ they are less practical for clinical or research use than plasma biomarkers. Yet, relationships between plasma pTau217 and clinical outcomes are less established. In LBD, higher plasma pTau217 is cross-sectionally associated with worse global cognition,^20^ with higher baseline levels also linked to future phenoconversion to PD in isolated rapid eye movement sleep disorder^21^ and progression of cognitive impairment in PD.^22^ Serial plasma pTau217 data are scarce in LBD, though one longitudinal study observed a faster 3-4 year pTau217 increase in participants with PD and worsening cognition.^22^ Emerging literature therefore supports plasma pTau217 as a potential biomarker of clinical progression in LBD, though longitudinal studies are needed and prior work is limited by reliance on global outcome measures, AD-derived or data-driven thresholds for abnormal pTau217, and inconsistently defined clinical contexts of use.

Therefore, the purpose of this study was to characterize the clinical utility of plasma pTau217 across three biomarker contexts of use in a deeply phenotyped cohort of clinically diagnosed LBD with longitudinal plasma and neuropsychological test data. First, we evaluated pTau217 as a prognostic biomarker by testing whether higher baseline levels were associated with faster 2-8-year cognitive and functional decline. We then evaluated pTau217 as a risk-stratification biomarker by applying an LBD-specific cut-point defining abnormal levels^17^ to determine if individuals with abnormal pTau217 demonstrated worse clinical trajectories and risk for diagnostic progression to MCI or dementia. Finally, we evaluated pTau217 as a monitoring biomarker by testing whether faster 2-6-year longitudinal increase was associated with worse cognitive and functional decline.

## Methods

### Study Cohort

Data from participants enrolled in the Iqbal Farrukh and Asad Jamal Stanford Alzheimer’s Disease Research Center (ADRC) from September 2015 to February 2023 were analyzed. All participants underwent annual clinical evaluations involving medical history, physical and neurologic examination, neuropsychological assessment, and biospecimen collection. ADRC study procedures and enrollment criteria are summarized in eMethods 1.1 and described elsewhere.^23^

For the current study, inclusion criteria were availability of plasma pTau217 data from ≥ 1 ADRC study visits and an ADRC clinical consensus diagnosis of LBD spectrum (LBD), AD spectrum (AD), or healthy control (HC) following the first study visit. The AD group was included for comparison given validated use of plasma pTau217 in AD cohorts.^14^ Exclusion criteria were clinical consensus diagnosis other than LBD, AD, or HC, missing clinical consensus diagnosis data across all ADRC study visits, and missing or out-of-range plasma pTau217 values (Figure 1).

**Figure 1.**
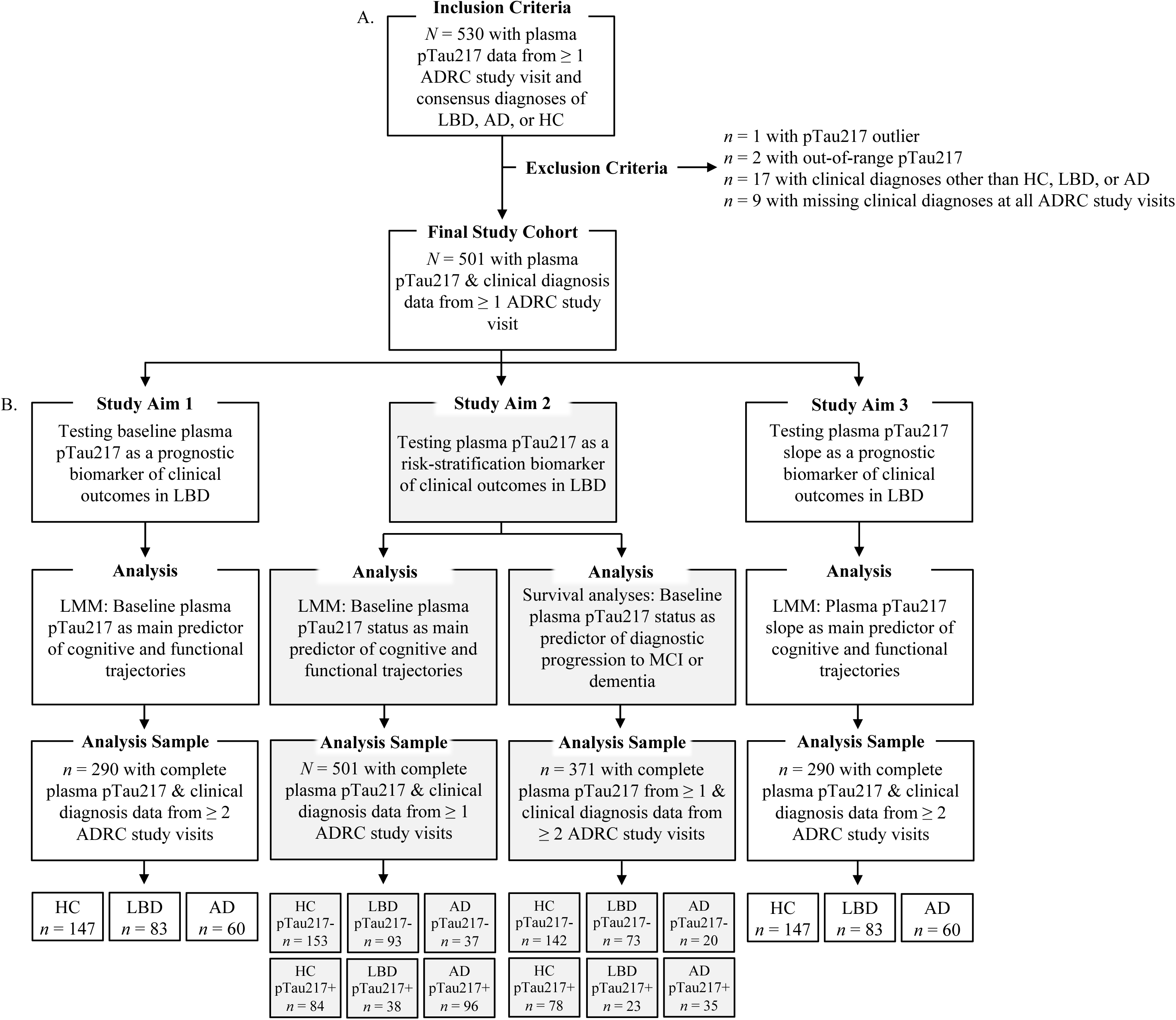
Cohort selection for overall study and longitudinal analyses evaluating each study aim. Panel (A.) illustrates inclusion and exclusion criteria for the overall study cohort. Panel (B.) details the analytic approach and sample size used to test each study aim. Aims 1 and 3 evaluated baseline plasma pTau217 and pTau217 slope as a prognostic and monitoring biomarker, respectively, in a sample of *n* = 290 using linear mixed-effects models. Aim 2 evaluated baseline plasma pTau217 status as a risk-stratification biomarker in a sample of *N* = 501 using separate linear mixed-effects models and in a sample of *n* = 371 using time-to-event analyses. AD = Alzheimer’s disease spectrum; ADRC = Stanford Alzheimer’s Disease Research Center; HC = Healthy control; LBD = Lewy body disease spectrum; LMM = Linear mixed effects models; pTau217+ = Baseline plasma pTau217 **>** 0.54pg/mL (abnormal); pTau217- = Baseline plasma pTau217 ≤ 0.54pg/mL (normal).

### Standard Protocol Approvals, Registrations, and Participant Consents

The Stanford University Institutional Review Board approved all study protocols. Participants or their legally authorized representative provided written informed consent in accordance with the Declaration of Helsinki. This study followed STROBE reporting guidelines.

### Clinical Diagnosis Groups

Clinical diagnoses were established during multidisciplinary consensus meetings using published criteria and available clinical data (eMethods 1.2). Participants were classified as neurologically healthy older adults (HC), AD (mild cognitive impairment [MCI] or dementia due to AD^24,25^), or LBD (PD with normal cognition [PDNC],^26^ PD with MCI,^27^ LBD with MCI/prodromal dementia with Lewy bodies,^28^ PD dementia,^29^ and dementia with Lewy bodies^30^). Baseline clinical diagnosis was determined at the first consensus meeting following ADRC study entry and included as a cross-sectional, categorical predictor in longitudinal analyses.

### Demographics and Covariates

Sex and education (highest level completed) from the first ADRC study visit and age at first cognitive or functional assessment were included in longitudinal models as covariates. Age and education were mean-centered.

### Plasma Collection and pTau217 Measurement

Whole blood was collected at each annual ADRC study visit and plasma was isolated using standard protocol (eMethods 1.3).

All ADRC participants with available cross-sectional and longitudinal plasma samples were tested for pTau217. Plasma pTau217 levels were quantified using the Quanterix Simoa^®^ HD-X platform and ALZpath Simoa® pTau217 V2 assay kit (#104371, Quanterix) at the Department of Psychiatry and Neurochemistry, University of Gothenburg. Assay workflow followed manufacturer and previously published guidelines (eMethods 1.3).^13^ Samples outside the quantifiable range for the limit of detection (0.0072-30.0 pg/mL) were excluded from analyses.

### Plasma pTau217 Analysis

Baseline and longitudinal plasma pTau217 values (eTable 1) were operationalized for each study aim. To evaluate plasma pTau217 as a prognostic biomarker, baseline pTau217 was included as a continuous predictor in linear mixed-effects models. Baseline pTau217 was defined as the first available measurement across ADRC study visits for each participant and was log_10_-transformed for interpretability (eFigure 1).

To evaluate plasma pTau217 as a risk-stratification biomarker, baseline pTau217 was dichotomized and included as baseline pTau217 status (abnormal versus normal) in separate linear mixed-effects and survival models. Since amyloid and tau burden are known to differ in LBD cohorts,^31^ we applied an amyloid PET-derived cut-point previously validated in participants with LBD and co-occurring ADNC to derive baseline pTau217 status. Raw pTau217 values > 0.54pg/mL were classified as abnormal (pTau217+) and values ≤ 0.54pg/mL as normal (pTau217-). Cut-point development details are summarized in eMethods 1.4 and described elsewhere.^17^

To evaluate plasma pTau217 as a monitoring biomarker, longitudinal pTau217 change was included as a continuous predictor in the same linear mixed-effects models as baseline pTau217. Longitudinal pTau217 change was examined as participant-specific annual slopes (pTau217 slope). Specifically, among individuals with 2-5 plasma pTau217 measurements across ADRC study visits (*n* = 290), individual linear regressions modeled log_10_-transformed pTau217 as a function of years since first pTau217 measurement. Mean (SD) number of pTau217 measurements was 2.6 (0.8) and mean (SD) duration between first and last measurement was 2.3 (1.4) years (range = 0.6-6.2 years; eFigure 2).

### Longitudinal Clinical Outcomes

The Clinical Dementia Rating-Sum of Boxes (CDR-SB; range = 0-18)^32^ and Montreal Cognitive Assessment (MoCA; range = 0-30)^33^ were administered at annual ADRC study visits, with scores analyzed over 2-8 years in separate linear mixed-effects models to test changes in daily functioning^34^ and global cognition, respectively (eMethods 1.5.1, 1.5.2; eResults 2.1; eTable 2).

At annual ADRC study visits, participants also completed neuropsychological tests from the National Alzheimer’s Coordinating Center Uniform Data Set (Version 3)^35^ and supplemental measures of memory, executive function, and visuospatial function.^23^ Participants in the LBD group on dopamine replacement therapy completed testing in the on-medication state as recommended.^27^ In the current study, baseline test scores (scores from first assessment after ADRC study entry) were obtained within six months of initial plasma collection.

Raw test scores were *z*-transformed using the baseline mean and standard deviation of participants with and without plasma pTau217 data classified as cognitively normal across all ADRC study visits (*n* = 301). Five cognitive indices were derived by averaging *z*-scores for tests measuring memory, executive function, visuospatial function, language, and processing speed (eMethods 1.5.3). This method yielded robust scores for distinct cognitive domains and handled missing data by allowing partial test score availability within each index.^36,37^ Tests were selected for each index based on previously published work^38,39^ and established domain structure of the ADRC Uniform Data Set neuropsychological battery.^35^ Higher index scores indicated better cognition. Scores from each index were analyzed in separate linear-mixed effects models to test change in domain-specific performance over 2-8 years (eResults 2.1, eTables 2-3).

### Statistical Analyses

All statistical analyses were performed in R version 4.1.1.^40^ Two-sided *p*-values of ≤ 0.05 were considered statistically significant.

### Descriptive Statistics

Non-normally distributed continuous variables were described using median (interquartile range). Categorical variables were described using counts (percentages). Group differences were assessed using Kruskal-Wallis or Mann-Whitney U tests for non-normally distributed continuous variables and Pearson’s chi-square tests for categorical variables. Post hoc pairwise comparisons between the three clinical diagnosis groups were conducted when appropriate using Wilcoxon rank-sum or chi-square tests with Bonferroni correction for multiple comparisons.

### Plasma pTau217 as a Prognostic and Monitoring Biomarker

A series of linear mixed-effects models evaluated baseline pTau217 and pTau217 slope as prognostic and monitoring biomarkers, respectively. Participants with ≥ 2 plasma pTau217 measurements (*n* = 290) were included because baseline pTau217 and pTau217 slope variables were analyzed as predictors simultaneously in each model to account for baseline level while testing effects of longitudinal change on clinical outcome trajectories.

Seven separate models were fit for longitudinal CDR-SB, MoCA, and five cognitive index scores using maximum likelihood estimation to include all available observations under the assumption of missing at random. Models included random intercepts and slopes with an unstructured covariance matrix to capture within-person variability in baseline performance and rate of change in clinical outcomes.

Time was defined as years since baseline assessment and modeled via linear or natural cubic spline specifications, allowing model flexibility for non-linear trends.^41^ Linear and spline models were compared using Akaike Information Criterion, Bayesian Information Criterion, and likelihood-ratio tests. Spline models were selected if they provided superior fit and improved interpretability (eTable 4). CDR-SB, MoCA, memory index, and language index scores were modeled using linear time. Executive function, visuospatial function, and processing speed index scores were modeled using natural cubic splines (eMethods 1.6.1).

Fixed effects included baseline pTau217, pTau217 slope, clinical diagnosis group, time, and covariates (age, sex, education). Baseline pTau217 and pTau217 slope were mean-centered within clinical diagnosis groups.

Interactions between baseline pTau217 or pTau217 slope, clinical diagnosis group, and time evaluated effects of pTau217 variables on clinical outcome trajectories within clinical diagnosis groups. Diagnosis-specific, simple slope estimates were derived using Wald tests, with Benjamini-Hochberg false discovery rate (FDR) corrections applied across three planned simple slope tests for baseline pTau217 and pTau217 slope separately.

### Plasma pTau217 Status as a Risk-Stratification Biomarker of Faster Cognitive and Functional Decline

In participants with ≥ 1 plasma pTau217 measurement (*N* = 501), separate linear mixed-effects models evaluated baseline plasma pTau217 status (abnormal versus normal) as a risk-stratification biomarker. Models were specified using the same methods described in the previous section, with baseline pTau217 status replacing baseline pTau217 and pTau217 slope as the predictor of interest. Model fit was similar across clinical outcomes (eMethods 1.6.3, eTable 4).

Interactions between baseline pTau217 status, clinical diagnosis group, and time evaluated effects of pTau217 status on clinical outcome trajectories in each clinical diagnosis group. Using two-sided Satterthwaite *t*-tests, simple slope contrasts compared annual clinical outcome change between abnormal and normal pTau217 status groups within each clinical diagnosis group, with FDR corrections applied across these three planned contrasts.

### Plasma pTau217 Status as a Risk-Stratification Biomarker of Diagnostic Progression to MCI or Dementia

Among participants without dementia at baseline and clinical diagnosis data at ≥ 2 ADRC study visits (*n* = 371), time-to-event analyses evaluated baseline plasma pTau217 status as a risk-stratification biomarker. Survival time was defined as years from first ADRC consensus diagnosis to first observed cognitive status conversion, with censoring at last ADRC study visit. In LBD and HC, conversion of cognitive status was defined as progression from cognitively unimpaired to MCI or MCI to dementia. In AD, conversion was defined as progression from MCI to dementia. Participants were stratified by baseline clinical diagnosis and pTau217 status: HC pTau217-, HC pTau217+, LBD pTau217-, LBD pTau217+, AD pTau217-, and AD pTau217+.

Kaplan-Meier curves estimated unadjusted time to conversion, with group differences evaluated using log-rank tests and Benjamini-Hochberg-adjusted pairwise comparisons. Five-year survival probabilities were estimated for interpretation. Additionally, Cox models adjusted for age and sex estimated hazard ratios relative to HC pTau217-, with one planned contrast comparing LBD pTau217+ and pTau217-groups. Proportional hazards assumptions were assessed using Schoenfeld residuals and were met. Sensitivity analyses stratifying Cox models by baseline cognitive status were performed (eMethods 1.6.4). Results did not change and baseline cognitive status was not analyzed in primary models (eResults 2.3; eTable 8).

To account for differences in baseline severity of LBD-related cognitive impairment, exploratory time-to-event analyses examined additional definitions of cognitive conversion. First, in cognitively unimpaired participants stratified by pTau217 status (*n* = 275; HC pTau217-, HC pTau217+, PDNC pTau217-, PDNC pTau217+), conversion was defined as progression to MCI or dementia, with one planned contrast comparing PDNC pTau217+ and PDNC pTau217-groups. Second, among individuals with MCI due to PD (MCI-PD) or prodromal dementia with Lewy bodies (MCI-LB) stratified by pTau217 status (*n* = 36; MCI-LB pTau217-, MCI-LB pTau217+, MCI-PD pTau217-, MCI-PD pTau217+), conversion was defined as progression to dementia. Sex was excluded as a covariate in Cox Models with MCI participants for violating proportional hazards assumptions.

### Data Availability

Anonymized data can be made available on request to qualified researchers in accordance with participant privacy regulations and contingent upon securing local approvals through appropriate data use agreements.

## Results

### Baseline Sample Characteristics

A total of 501 participants classified as LBD (*n* = 133), AD (*n* = 131), or HC (*n* = 237) were included. Table 1 describes baseline sample characteristics stratified by clinical diagnosis group.

**Table 1.** Baseline characteristics for demographics, plasma pTau217, and clinical outcomes by clinical diagnosis (*N* = 501)

| | HC<br>($n = 237$) | LBD<br>($n = 131$) | AD<br>($n = 133$) | Test<br>Statistic | $p$-value |
| --- | --- | --- | --- | --- | --- |
| Age *Median (IQR)* | 72 (67, 77) | 70 (64, 75) | 73 (65, 78) | 8.13<sup>a</sup> | 0.02 |
| Education *Median (IQR)* | 18 (16, 18) | 18 (16, 18) | 16 (14, 18) | 5.96<sup>a</sup> | 0.05 |
| Sex $n$ (%)<sup>c</sup> | | | | 25.45<sup>b</sup> | < 0.001 |
| Female | 150 (63.29) | 51 (38.93) | 57 (42.86) |  |  |
| Male | 87 (36.71) | 80 (61.07) | 76 (57.14) |  |  |
| Ethnicity $n$ (%) | | | | 5.54<sup>b</sup> | 0.06 |
| Non-Hispanic/Latino | 205 (86.50) | 123 (93.89) | 114 (85.71) |  |  |
| Hispanic/Latino | 32 (13.50) | 8 (6.11) | 19 (14.29) |  |  |
| Race $n$ (%) | | | | 15.48<sup>b</sup> | 0.12 |
| White | 207 (87.34) | 124 (94.66) | 115 (86.47) |  |  |
| Black/African American | 2 (0.84) | 1 (0.76) | 4 (3.01) |  |  |
| American Indian/Alaska Native | 1 (0.42) | 0 (0.00) | 2 (1.50) |  |  |
| Native Hawaiian/Pacific Islander | 0 (0.00) | 0 (0.00) | 1 (0.75) |  |  |
| Asian | 25 (10.55) | 6 (4.58) | 9 (6.77) |  |  |
| Other | 2 (0.84) | 0 (0.00) | 2 (1.5) |  |  |
| *APOE* $\epsilon 4$ Carrier Status $n$ (%)<sup>c</sup> | | | | 39.98<sup>b</sup> | < 0.001 |
| No | 169 (71.61) | 92 (70.23) | 52 (40.31) |  |  |
| Yes | 67 (28.39) | 39 (29.77) | 77 (59.69) |  |  |
| pTau217 Status $n$ (%)<sup>d</sup> | | | | 61.96<sup>a</sup> | < 0.001 |
| Normal | 153 (64.56) | 93 (70.99) | 37 (27.82) |  |  |
| Abnormal | 84 (35.44) | 38 (29.01) | 96 (72.18) |  |  |
| Baseline pTau217 *Median (IQR)*<sup>d</sup> | 0.38 (0.28, 0.64) | 0.37 (0.29, 0.57) | 0.92 (0.52, 1.61) | 108.95<sup>a</sup> | < 0.001 |
| pTau217 Slope *Median (IQR)*<sup>d,e</sup> | 0.02 (-0.03, 0.09) | 0.04 (-0.01, 0.10) | 0.05 (-0.06, 0.18) | 5.45<sup>a</sup> | 0.066 |
| CDR-SB *Median (IQR)* | 0.0 (0.0, 0.0) | 0.5 (0.0, 1.0) | 2.5 (1.0, 4.5) | 213.56<sup>a</sup> | < 0.001 |
| MoCA *Median (IQR)* | 27 (26, 28) | 26 (23, 28) | 20 (15, 23) | 168.31<sup>a</sup> | < 0.001 |
| Cognitive Indices *Median (IQR)* |  |  |  |  |  |
| Memory | 0.06 (-0.42, 0.48) | -0.32 (-1.20, 0.07) | -2.71 (-3.66, -1.46) | 214.82<sup>a</sup> | < 0.001 |
| Executive Function | 0.01 (-0.46, 0.40) | -0.45 (-1.02, 0.17) | -1.18 (-2.00, -0.44) | 103.95<sup>a</sup> | < 0.001 |
| Visuospatial Function | 0.10 (-0.40, 0.48) | -0.39 (-1.06, 0.21) | -0.61 (-1.62, -0.01) | 60.20<sup>a</sup> | < 0.001 |
| Language | 0.09 (-0.33, 0.45) | -0.45 (-0.99, 0.17) | -1.07 (-1.90, -0.45) | 129.21<sup>a</sup> | < 0.001 |
| Processing Speed | 0.00 (-0.53, 0.40) | -0.23 (-0.97, 0.23) | -0.92 (-2.08, -0.37) | 64.41<sup>a</sup> | < 0.001 |

**Note.** Abbreviations: AD = Alzheimer's disease spectrum; HC = Healthy controls; IQR = Interquartile range; LBD = Lewy body disease spectrum; Abnormal = Baseline plasma pTau217 > 0.54pg/mL; Normal = Baseline plasma pTau217 ≤ 0.54pg/mL. Baseline values for each clinical outcome were defined as the first assessment across ADRC study visits and exclude participants with missing data at all study visits. Baseline sample sizes for clinical outcomes varied with regard to missingness, and descriptives were calculated using available data. Sample sizes contributing to each clinical outcome over time are provided in eTable 2.
<sup>a</sup>Kruskal-Wallis tests.
<sup>b</sup>Pearson's $\chi^2$ tests.
<sup>c</sup>Five participants (AD, $n = 4$ ; HC, $n = 1$ ) were missing *APOE* $\epsilon 4$ carrier status data.
<sup>d</sup>Descriptive statistics reflect raw pTau217 values and inferential statistics used log<sub>10</sub>-transformed pTau217 values.
<sup>e</sup>Sample included participants with $\geq 2$ plasma measurements ( $n = 290$ ).

Age, sex, and *APOE* ε4 carrier status significantly differed between groups. LBD was younger (*p* = 0.02) and had more males (*p* < 0.001) compared to HC. LBD had fewer *APOE* ε4 carriers compared to AD (*p* < 0.001). Education, race, and ethnicity did not significantly differ between groups.

Baseline pTau217 significantly differed between groups. LBD had lower baseline pTau217 than AD (*p* < 0.001), though did not significantly differ from HC (*p* = 0.78). Baseline pTau217 status also significantly differed between groups. LBD had fewer participants with abnormal pTau217 compared to AD (*p* < 0.001), though did not significantly differ from HC (*p* = 0.76). Between-group differences in pTau217 slope were not significant (*p* = 0.07).

CDR-SB and MoCA scores significantly differed between groups. LBD had higher CDR-SB scores than HC (*p* < 0.001), but lower CDR-SB scores than AD (*p* < 0.001). LBD also had lower MoCA scores than HC (*p* = 0.009), but higher MoCA scores than AD (*p* < 0.001). All baseline cognitive indices significantly differed between groups (all *p* < 0.001). LBD performed worse than HC (all *p* ≤ 0.008) but better than AD (all *p* < 0.001) across all indices except visuospatial function (*p* = 0.09).

### Baseline Plasma pTau217 as a Prognostic Biomarker

eFigure 3 shows observed and model-estimated effects of baseline pTau217 on clinical outcome trajectories. In LBD, higher baseline pTau217 was associated with faster increase in CDR-SB (*β* = 0.30, *p* < 0.001) and decline in MoCA (*β* = −0.35, *p* = 0.002). Higher baseline pTau217 was also associated with faster declines in memory (*β* = −0.06, *p* = 0.015), executive function (*β* = −1.47, *p* = 0.001), and visuospatial function (*β* = −2.15, *p* = 0.028) indices (Table 2). Sensitivity analyses of non-linear baseline pTau217 effects (eMethods 1.6.2) did not materially alter LBD findings (eResults 2.2, eTables 5-6).

**Table 2.**
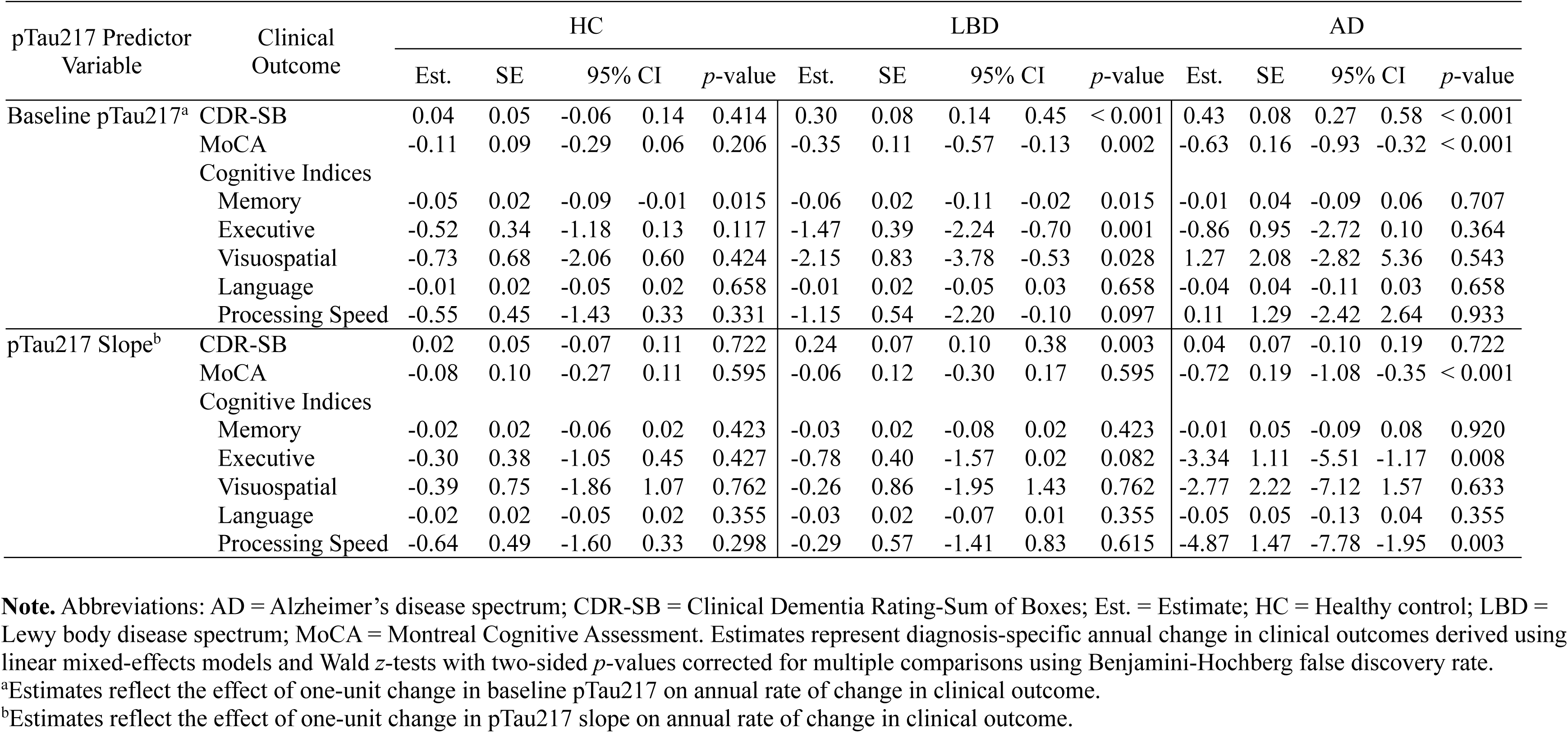
Effects of baseline plasma pTau217 and plasma pTau217 slope on longitudinal cognitive and functional trajectories (*n* = 290)

In AD, higher baseline pTau217 was associated with faster increase in CDR-SB (*β* = 0.43, *p* < 0.001) and decline in MoCA (*β* = −0.63, *p* < 0.001), though was not significantly associated with longitudinal cognitive index change. In HC, higher baseline pTau217 was associated with faster memory index decline (*β* = −0.05, *p* = 0.015; Table 2).

### Plasma pTau217 Status as a Risk-Stratification Biomarker of Faster Cognitive and Functional Decline

Figure 2 displays observed and model-estimated clinical outcome trajectories stratified by baseline pTau217 status. In LBD, participants with abnormal pTau217 demonstrated faster increase in CDR-SB (**Δ** = 0.85 points/year, *p* < 0.001) and decline in MoCA (**Δ** = −0.87 points/year, *p* = 0.001) compared to those with normal pTau217. Additionally, participants with abnormal pTau217 demonstrated faster declines in memory (**Δ** = −0.16 points/year, *p* = 0.006), executive function (**Δ** = −0.22 points/year, *p* = 0.011), and visuospatial function (**Δ** = −0.43 points/year, *p* = 0.018) indices compared to those with normal pTau217 (Table 3).

**Figure 2.**
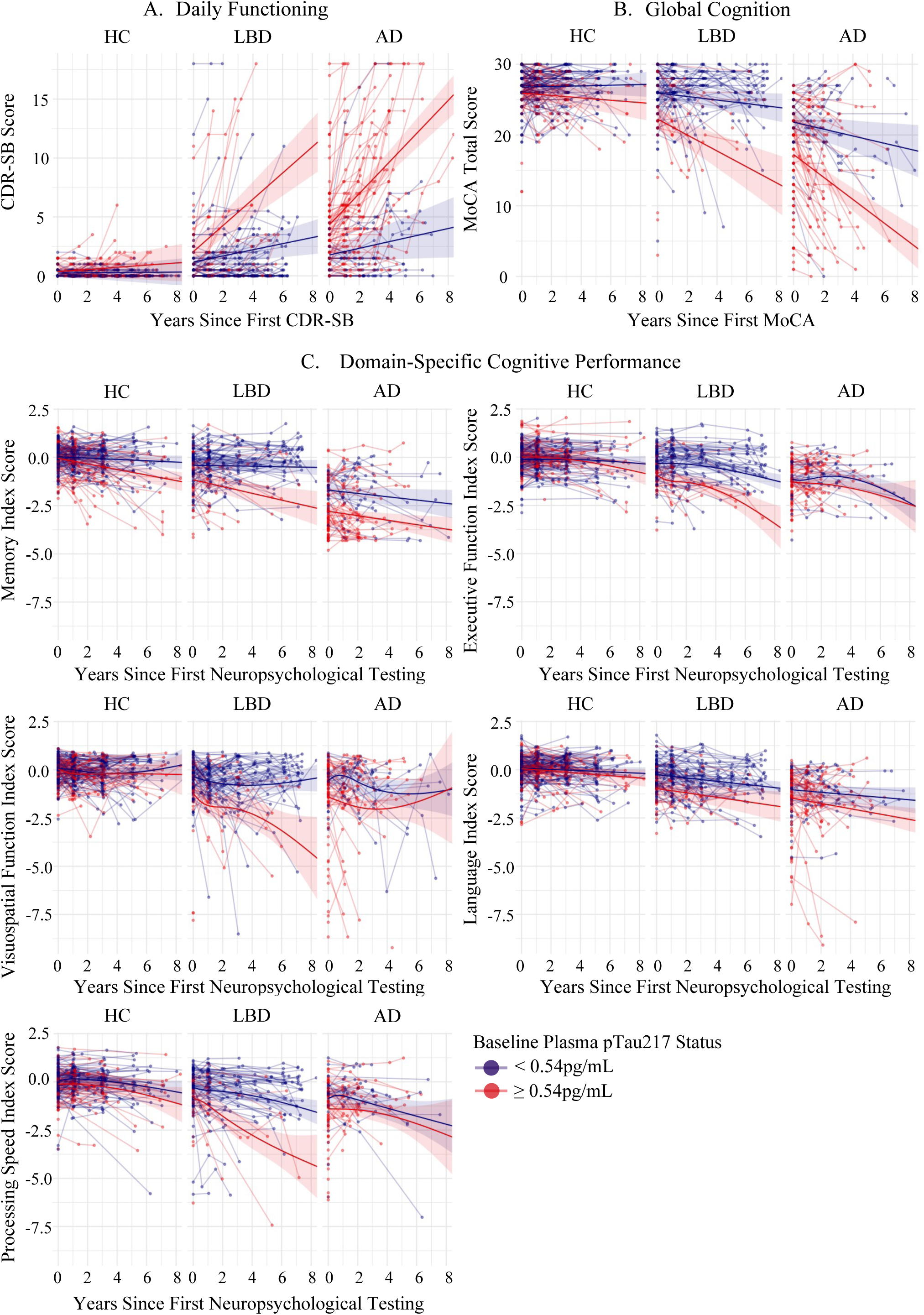
Effect of baseline plasma pTau217 status on clinical outcome trajectories. Observed participant (thin lines) and model-estimated (bold lines with 95% confidence interval shading) trajectories are shown for each clinical diagnosis group (HC, LBD, AD) stratified by baseline plasma pTau217 status for: (A.) CDR-SB, (B.) MoCA, and (C.) five cognitive index scores over 2-8 years. Points represent individual clinical outcome observations. Red lines and points represent participants with abnormal baseline pTau217 (> 0.54pg/mL) and blue lines and points represent participants with normal baseline pTau217 (≤ 0.54pg/mL). AD = Alzheimer’s disease spectrum; CDR-SB = Clinical Dementia Rating-Sum of Boxes; HC = Healthy control; LBD = Lewy body disease spectrum; MoCA = Montreal Cognitive Assessment.

**Table 3.** Estimated annual clinical outcome change stratified by baseline plasma pTau217 status and clinical diagnosis (*N* = 501)

| Clinical Diagnosis Group | Clinical Outcome | Normal pTau217<sup>a</sup> | Abnormal pTau217<sup>a</sup> | SE<sup>b</sup> | 95% CI<sup>b</sup> |  | *t*-value<sup>b</sup> | *p*-value<sup>b,c</sup> |
| --- | --- | --- | --- | --- | --- | --- | --- | --- |
| HC | CDR-SB | 0.011 | 0.091 | 0.099 | -0.114 | 0.274 | 0.815 | 0.416 |
|  | MoCA | 0.046 | -0.170 | 0.165 | -0.541 | 0.110 | -1.306 | 0.193 |
|  | Cognitive Indices |  |  |  |  |  |  |  |
|  | Memory | -0.035 | -0.144 | 0.034 | -0.176 | -0.041 | -3.171 | 0.005 |
|  | Executive Function | -0.031 | -0.109 | 0.440 | -0.171 | 0.016 | -1.628 | 0.157 |
|  | Visuospatial Function | 0.036 | -0.032 | 0.840 | -0.247 | 0.111 | -0.747 | 0.574 |
|  | Language | -0.032 | -0.060 | 0.030 | -0.088 | 0.032 | -0.918 | 0.539 |
|  | Processing Speed | -0.078 | -0.136 | 0.673 | -0.202 | 0.085 | -0.800 | 0.636 |
| LBD | CDR-SB | 0.259 | 1.110 | 0.150 | 0.557 | 1.145 | 5.688 | < 0.001 |
|  | MoCA | -0.250 | -1.123 | 0.256 | -1.378 | -0.367 | -3.404 | 0.001 |
|  | Cognitive Indices |  |  |  |  |  |  |  |
|  | Memory | -0.014 | -0.176 | 0.055 | -0.271 | -0.053 | -2.934 | 0.006 |
|  | Executive Function | -0.121 | -0.343 | 0.702 | -0.371 | -0.073 | -2.923 | 0.011 |
|  | Visuospatial Function | 0.004 | -0.422 | 1.423 | -0.728 | -0.123 | -2.762 | 0.018 |
|  | Language | -0.083 | -0.112 | 0.049 | -0.125 | 0.069 | -0.574 | 0.566 |
|  | Processing Speed | -0.161 | -0.412 | 1.057 | -0.476 | -0.026 | -2.197 | 0.086 |
| AD | CDR-SB | 0.315 | 1.334 | 0.155 | 0.713 | 1.324 | 6.561 | < 0.001 |
|  | MoCA | -0.514 | -1.612 | 0.260 | -1.610 | -0.586 | -4.222 | < 0.001 |
|  | Cognitive Indices |  |  |  |  |  |  |  |
|  | Memory | -0.087 | -0.117 | 0.058 | -0.143 | 0.084 | -0.511 | 0.610 |
|  | Executive Function | -0.192 | -0.162 | 1.007 | -0.184 | 0.244 | 0.276 | 0.783 |
|  | Visuospatial Function | -0.030 | 0.095 | 2.054 | -0.312 | 0.562 | 0.562 | 0.574 |
|  | Language | -0.067 | -0.141 | 0.054 | -0.181 | 0.033 | -1.356 | 0.529 |
|  | Processing Speed | -0.166 | -0.176 | 1.465 | -0.322 | 0.301 | -0.065 | 0.948 |

**Note.** Abbreviations: AD = Alzheimer's disease spectrum; CDR-SB = Clinical Dementia Rating-Sum of Boxes; Est. = Estimate; HC = healthy control; LBD = Lewy body disease spectrum; MoCA = Montreal Cognitive Assessment. Table 3 estimates are reported to three decimal places to clarify the calculation of the differences reported in the Results. Abnormal pTau217 was defined as baseline plasma pTau217 > 0.54pg/mL and Normal pTau217 as baseline plasma pTau217 ≤ 0.54pg/mL.
<sup>a</sup>Estimates represent model-estimated annual change in clinical outcome within abnormal and normal pTau217 status groups for each clinical diagnosis group.
<sup>b</sup>SE, 95% CIs, *t*-value, and *p*-value correspond to Satterthwaite *t*-tests evaluating differences in annual clinical outcome change between abnormal and normal pTau217 status groups.
<sup>c</sup>Corrected for multiple comparisons using Benjamini-Hochberg false discovery rate.

In AD, participants with abnormal baseline pTau217 demonstrated faster increase in CDR-SB (**Δ** = 1.03 points/year, *p* < 0.001) and decline in MoCA (**Δ** = −1.10 points/year, *p* < 0.001) compared to those with normal levels, though cognitive index trajectories did not significantly differ by pTau217 status. In HC, participants with abnormal pTau217 demonstrated faster memory index decline (**Δ** = −0.11 points/year, *p* = 0.005) compared to those with normal levels (Table 3).

### Plasma pTau217 Status as a Risk-Stratification Biomarker of Diagnostic Progression to MCI or Dementia

Time to conversion significantly differed between groups stratified by clinical diagnosis and pTau217 status (log-rank χ² = 68.4[5]; *p* < 0.001). LBD pTau217+ demonstrated the highest conversion risk followed by AD pTau217+. LBD pTau217- and AD pTau217- showed overlapping and intermediate risk trajectories, and HC pTau217+ had the lowest conversion risk (Figure 3). In adjusted Cox models (*n* = 371; 80 events), all groups had significantly higher conversion risk compared to HC pTau217- (Table 4). The planned contrast found LBD pTau217+ had a three-fold higher conversion risk compared to LBD pTau217- (HR = 3.41; 95% CI = [1.60, 7.28]; *p* = 0.001).

**Figure 3.**
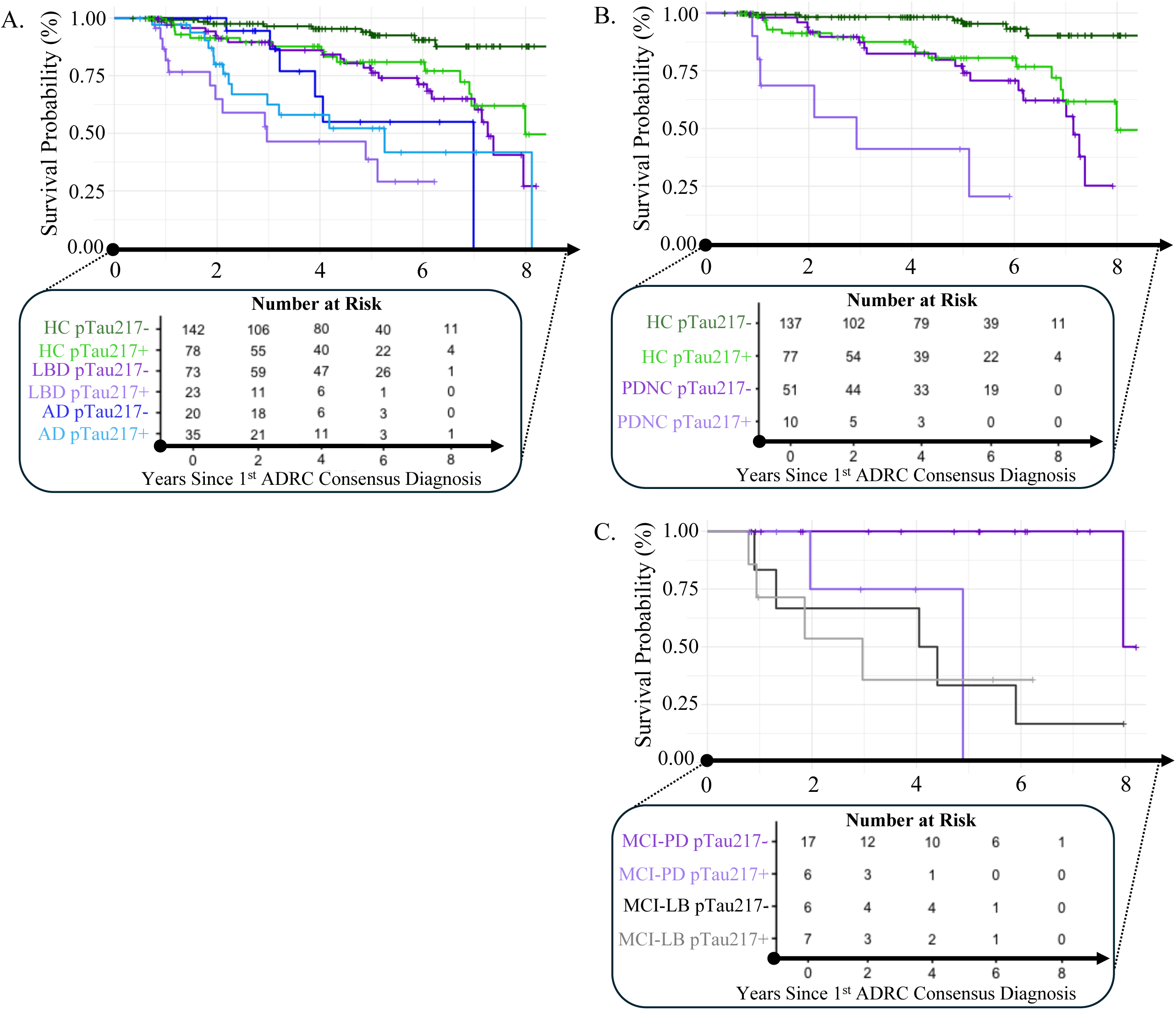
Effect of baseline plasma pTau217 status on diagnostic progression to MCI or dementia. Unadjusted Kaplan-Meier survival curves depicting time to conversion to MCI or dementia for participants grouped by (A.) clinical diagnosis (HC, AD, LBD) and pTau217 status (pTau217+, pTau217-), (B.) cognitively unimpaired diagnosis (HC and PDNC) and pTau217 status, and (C.) MCI diagnosis (LB-MCI and PD-MCI) and pTau217 status. The x-axis shows years from first clinical diagnosis to observed cognitive conversion or censoring. The y-axis shows survival probability defined as proportion of participants who did not convert to MCI or dementia at a given timepoint, with corresponding frequencies shown below the x-axis (Number at Risk). Tick marks on survival curves represent censored observations. AD = Alzheimer’s disease spectrum; CDR-SB = Clinical Dementia Rating-Sum of Boxes; HC = healthy control; LBD = Lewy body disease spectrum; MCI = Mild cognitive impairment; MCI-LB = Mild cognitive impairment due to Lewy body disease (prodromal dementia with Lewy bodies); MCI-PD = Mild cognitive impairment due to Parkinson’s disease; PDNC = Parkinson’s disease with normal cognition; pTau217+ = > 0.54pg/mL (abnormal); pTau217- = ≤ 0.54pg/mL (normal).

**Table 4.** Cox models for diagnostic progression to MCI or dementia stratified by baseline plasma pTau217 status and clinical diagnosis.

| Conversion Event | Model Predictor | Hazard Ratio | 95% CI |  | p-value |
| --- | --- | --- | --- | --- | --- |
| MCI or Dementia<br>(*n* = 371)<sup>a,b</sup> | HC pTau217- | 1.00 | - | - | - |
|  | HC pTau217+ | 3.26 | 1.43 | 7.41 | 0.005 |
|  | AD pTau217- | 6.44 | 2.23 | 18.60 | 0.001 |
|  | AD pTau217+ | 9.78 | 4.20 | 22.77 | < 0.001 |
|  | LBD pTau217- | 5.18 | 2.36 | 11.39 | < 0.001 |
|  | LBD pTau217+ | 17.88 | 7.16 | 44.64 | < 0.001 |
|  | Age | 1.03 | 0.99 | 1.06 | 0.111 |
|  | Sex | 1.00 | 0.62 | 1.60 | 0.903 |
| MCI or Dementia<br>(*n* = 275)<sup>c</sup> | HC pTau217- | 0.11 | 0.04 | 0.28 | < 0.001 |
|  | HC pTau217+ | 0.51 | 0.25 | 1.07 | 0.077 |
|  | PDNC pTau217- | 1.00 | - | - | - |
|  | PDNC pTau217+ | 4.25 | 1.50 | 12.02 | 0.006 |
|  | Age | 1.05 | 1.00 | 1.09 | 0.042 |
|  | Sex | 0.87 | 0.46 | 1.65 | 0.675 |
| Dementia<br>(*n* = 36)<sup>d,e</sup> | MCI-PD pTau217- | 1.00 | - | - | - |
|  | MCI-PD pTau217+ | 22.21 | 1.44 | 343.02 | 0.026 |
|  | MCI-LB pTau217- | 19.20 | 2.00 | 184.26 | 0.010 |
|  | MCI-LB pTau217+ | 61.29 | 2.74 | 1367.04 | 0.009 |
|  | Age | 0.92 | 0.81 | 1.09 | 0.186 |

**Note.** Abbreviations: AD = Alzheimer's disease spectrum; HC = Healthy control; LBD = Lewy body disease spectrum; pTau217+ = Baseline plasma pTau217 ≥ 0.54pg/mL (abnormal); pTau217- = Baseline plasma pTau217 < 0.54pg/mL (normal). Hazard ratios were derived from multivariable Cox models adjusted for age and sex unless otherwise noted. Hazard ratios >1 indicate greater hazard of diagnostic progression relative to the reference group.
<sup>a</sup>Results reported relative to HC pTau217-.
<sup>b</sup>Conversion was defined as progression from cognitively unimpaired to MCI or MCI to dementia in HC and LBD, and progression from MCI to dementia in AD.
<sup>c</sup>Results reported relative to PDNC pTau217-.
<sup>d</sup>Results reported relative to MCI-PD pTau217-.
<sup>e</sup>Sex was removed as a covariate due to violating proportional hazards assumptions ( $\chi^2 = 1.12[3]$ , $p = 0.0045$ ).

Among cognitively unimpaired participants, time to conversion significantly differed by plasma pTau217 status (log-rank χ² = 57.2[3]; *p* < 0.001, eTable 7). PDNC pTau217+ had the highest conversion risk, PDNC pTau217- and HC pTau217+ showed overlapping and intermediate risk trajectories, and HC pTau217- had the lowest risk. (Figure 3). In adjusted Cox models (*n* = 275; 46 events), all groups had a significantly higher conversion risk compared to HC pTau217- (all *p* < 0.001). The planned contrast found PDNC pTau217+ had a four-fold higher conversion risk (HR = 4.25; 95% CI = [1.50, 12.02]; *p* = 0.006) compared to PDNC pTau217- (Table 4).

Among participants with MCI, time to conversion significantly differed by pTau217 status (log-rank χ² = 11.8[3]; *p* < 0.008, eTable 7). MCI-LB pTau217+ had the highest conversion risk, MCI-PD pTau217+ and MCI-LB pTau217- showed overlapping and intermediate risk trajectories, and MCI-PD pTau217- had the lowest conversion risk. (Figure 3). In adjusted Cox models (*n* = 36; 12 events; Table 4), MCI-LB pTau217-, MCI-LB pTau217+, and MCI-PD pTau217+ had significantly greater conversion risks compared to MCI-PD pTau217- (all *p* ≤ 0.026).

### Longitudinal Plasma pTau217 Change as a Monitoring Biomarker

eFigure 4 shows observed and model-estimated effects of pTau217 slope on clinical outcome trajectories. In LBD, a steeper pTau217 slope was associated with faster increase in CDR-SB (*β* = 0.24, *p* = 0.003). Plasma pTau217 slope was not significantly associated with longitudinal MoCA or cognitive index change (Table 2).

In AD, a steeper pTau217 slope was associated with faster declines in MoCA (*β* = −0.72, *p* < 0.001), executive function index (*β* = −3.34, *p* = 0.008), and processing speed index (*β* = −4.87, *p* = 0.003). In HC, pTau217 slope was not significantly associated with longitudinal clinical outcome change (Table 2).

## Discussion

This study characterized biomarker contexts of use for plasma pTau217 in LBD cohorts with co-occurring ADNC and varying severities of cognitive impairment. Specifically, higher baseline pTau217 was associated with faster cognitive and functional decline, demonstrating prognostic utility and suggesting a single blood draw yields meaningful information about clinical decline. Applying an LBD-specific cut-point also demonstrated risk-stratification value such that participants with abnormal baseline pTau217 had worse clinical trajectories characterized by a 0.8 point/year faster CDR-SB increase, 0.87 point/year faster MoCA decline, and three-fold greater risk for diagnostic progression to MCI or dementia within five years. Finally, faster annual increase in plasma pTau217 was associated with faster functional decline, suggesting repeat measurement may be relevant for monitoring changes in daily functioning related to cognitive decline or underlying, co-occurring disease processes.

### Baseline Plasma pTau217 as a Prognostic Biomarker

Prior cross-sectional work has demonstrated associations between pTau217 and ADNC,^15,16^ worse global cognition, and greater functional impairment^18,20^ in LBD-spectrum cohorts. Using similar outcomes, we show that a single baseline pTau217 measurement was also associated with declines up to eight years later, supporting prognostic utility in LBD. In the context of mixed pathology, baseline pTau217 may be particularly sensitive to global cognitive decline relative to other blood-based biomarkers. For instance, pTau181 is inconsistently shown to be associated with longitudinal global cognitive change.^39,42^ Additionally, while biomarkers such as neurofilament light chain protein and glial fibrillary associated protein have been associated with longitudinal declines in global cognition in LBD, they are not specific to ADNC.^43,44^ More research is needed comparing prognostic performance between plasma biomarkers.

Beyond global cognition, higher baseline plasma pTau217 was associated with faster domain-specific declines in memory, executive function, and visuospatial function in LBD. This parallels findings in symptomatic AD-spectrum cohorts where higher baseline pTau217 was associated with worse performance on individual measures of memory, language, executive function, and visuospatial function.^45^ Overlapping patterns of pTau217-related cognitive decline between LBD and AD suggest baseline pTau217 is related to cognitive heterogeneity in LBD linked to mixed pathology, and can potentially serve as a prognostic clinical tool or aid clinical trial enrichment.

Higher baseline plasma pTau217 was associated with longitudinal decline in global, but not domain-specific cognition in AD. This differs from preclinical and early AD research reporting associations between baseline pTau217 and task-specific decline.^46^ Since our AD cohort included participants with MCI and dementia, null findings may reflect limited within-person variability in clinical outcomes due to floor effects, particularly since higher baseline pTau217 was associated with memory decline in HC. Null domain-specific effects may also reflect restricted plasma pTau217 variability in symptomatic AD. Soluble phosphorylated tau increases early due to amyloid-β deposition. However, as amyloid-β burden plateaus and cortical tau accumulates, plasma pTau217 may better measure widespread tau burden and overall disease severity rather than amyloid-β-related changes.^47,48^ This suggests baseline pTau217 offers less incremental prognostic value for nuanced cognitive decline as AD pathology progresses. Baseline pTau217 may therefore be more informative in the context of lower cortical amyloid-β or tau burden, underscoring its utility in early LBD and mixed pathology groups.

### Baseline Plasma pTau217 Status as a Risk-Stratification Biomarker

Using an LBD-specific cut-point to define abnormal baseline plasma pTau217, we found that individuals with abnormal levels demonstrated faster annual declines in daily functioning and cognitive domains linked to future dementia onset. For instance, individuals with abnormal pTau217 had the fastest annual decline in visuospatial function out of all cognitive indices. This 0.4 SD/year rate of decline translates to clinically meaningful impairment after a few years given that scores between 1-1.5 SD below someone’s baseline typically represent a significant decline on neuropsychological tasks. This aligns with established relationships between early visuospatial impairment and dementia risk in LBD, and suggests abnormal baseline pTau217 is associated with at-risk clinical trajectories.^49^

In LBD, individuals with abnormal pTau217 also had the highest risk for progressing diagnostically to MCI or dementia, with 63% converting to dementia after five years compared to 24% of individuals with normal pTau217. This exceeded progression to dementia in AD with abnormal pTau217 (48%) and underscores the impact of concurrent ADNC in LBD. In PDNC, individuals with abnormal pTau217 also had the highest risk of diagnostic progression to MCI or dementia, supporting plasma pTau217 as a risk-stratification tool in early disease stages.

While exploratory analyses found that individuals with MCI-LB and abnormal pTau217 had higher risk for diagnostic progression to dementia than MCI-PD, sample sizes were small and findings should be interpreted cautiously. Replication in larger, independent cohorts is needed to differentiate dementia risk between MCI-LB and MCI-PD with abnormal pTau217.

### Longitudinal Plasma pTau217 as a Monitoring Biomarker

Consistent with emerging data, we found that a faster 2-6-year annual increase in pTau217 was associated with faster functional decline. This finding supports serial pTau217 measurement as a potential tool for tracking daily functioning change in LBD mixed pathology cohorts, a clinical population characterized by faster functional decline relative to AD.^50^ Additionally, while daily functioning is an important outcome for clinical trials, it is challenging to measure objectively. Longitudinal plasma pTau217 measured over short intervals may provide a scalable way to track long-term functional decline in clinically heterogeneous LBD populations.

Conversely, longitudinal pTau217 change was not associated with cognitive decline in LBD. Limited plasma sampling over short follow-up intervals and modest annual biomarker change may have reduced power to detect subtle biomarker-cognition associations. Also, modest pTau217 annual change may have contributed less variance to cognitive trajectories than baseline pTau217 and other drivers of cognitive decline like α-synuclein. Prior work in the same cohort found associations between faster three-year increases in plasma pTau181 and subsequent memory decline, suggesting pTau217 may be less clinically informative over similar intervals. ^39,51^ Studies with more frequent plasma sampling are needed to clarify clinical utility of longitudinal pTau217 change in LBD.

### Strengths and Limitations

Our study has several strengths. We leveraged a deeply phenotyped LBD cohort with longitudinal plasma pTau217 and up to eight years of clinical outcome follow-up. Domain-specific cognitive indices also allowed for nuanced characterization of associations between plasma pTau217 and symptom progression in LBD with co-occurring ADNC. Additionally, application of an amyloid PET-derived, LBD-specific cut-point improved clinical interpretability of findings. In addition to cut-point analyses, future studies in larger cohorts should examine non-linear relationships (threshold or plateau effects) between pTau217 and clinical progression that may help refine clinically meaningful biomarker thresholds in LBD.

This study also has limitations. First, diagnosis groups were clinically defined to reflect our study focus and the ADRC cohort design which utilized biomarker data for some, but not all, participants with consensus diagnoses. This can introduce diagnostic ambiguity or misclassification in mixed pathology cohorts, and abnormal plasma pTau217 is considered a proxy of ADNC in our sample. Future multimodal studies should therefore compare prognostic and monitoring value of plasma pTau217 to PET, CSF, or genetic biomarkers of ADNC. Additionally, mortality was not modeled as a competing risk in survival analyses due to limited data availability, and risk estimates reflect observed clinical progression among enrolled participants. Participants were also recruited from a single-site research cohort with limited racial or ethnic diversity, underscoring the need for additional longitudinal analyses of plasma pTau217 in more diverse LBD cohorts.

## Conclusion

In LBD, higher baseline levels and faster longitudinal increases in plasma pTau217 were associated with faster cognitive and functional decline. Abnormal baseline pTau217 was also associated with a three-fold greater risk of diagnostic progression to MCI or dementia. These findings support plasma pTau217 as a clinically meaningful prognostic, risk-stratification, and monitoring biomarker in LBD with co-occurring ADNC.

## Supporting information

Supplemental materials including: eMethods, eResults, eReferences, eTables, and eFigures

## Acknowledgements

This work was supported by the National Institutes of Health under award numbers P30 AG066515. This manuscript is the result of funding in whole or in part by the National Institutes of Health (NIH). It is subject to the NIH Public Access Policy. Through acceptance of this federal funding, NIH has been given a right to make this manuscript publicly available in PubMed Central upon the Official Date of Publication, as defined by NIH. The authors thank the Stanford Alzheimer’s Disease Research Center (ADRC) participants and staff for their contributions. Plasma pTau217 assays were graciously performed at the University of Gothenburg.

## Author Contributions

**S. A. Lorkiewicz:** drafting/revision of manuscript for content; analysis or interpretation of data; study concept or design. **C. Abdelnour:** drafting/revision of manuscript for content; analysis or interpretation of data; study concept or design. **M. L. Bolen:** drafting/revision of manuscript for content; analysis or interpretation of data. **A. M. Smith:** drafting/revision of manuscript for content; major role in acquisition of data. **M. Shahid-Besanti:** drafting/revision of manuscript for content; major role in acquisition of data. **D. Hemachandra:** drafting/revision of manuscript for content; analysis or interpretation of data. **E. Müller-Oehring:** drafting/revision of manuscript for content; analysis or interpretation of data. **N. Siddiqui:** Drafting/revision of manuscript for content. **L. Montoliu-Gaya:** drafting/revision of manuscript for content; major role in acquisition of data. **B. Arslan:** drafting/revision of manuscript for content; major role in acquisition of data. **N. J. Ashton:** drafting/revision of manuscript for content; major role in acquisition of data. **E. N. Wilson:** drafting/revision of manuscript for content; analysis or interpretation of data. **L. Tian:** drafting/revision of manuscript for content; analysis or interpretation of data. **K. I. Andreasson:** drafting/revision of manuscript for content; study concept or design. **E. C. Mormino:** drafting/revision of manuscript for content; study concept or design. **V. W. Henderson:** drafting/revision of manuscript for content; study concept or design. **H. Zetterberg:** drafting/revision of manuscript for content; major role in acquisition of data. **K. L. Poston:** drafting/revision of manuscript for content; study concept or design; supervision of the study; analysis or interpretation of data.

## References

1. Zarkali A, Bartl M, Fox NC, Tan LCS, Mollenhauer B, Weil RS. Diagnostic and other biomarkers of dementia with Lewy bodies: from research to clinical settings. Lancet Neurol. 2025;24(12):1053–1065. doi:10.1016/S1474-4422(25)00314-X

2. Scholz SW, Okubadejo NU, Prakash P, Liddelow SA, Ryten M, Halliday GM. Advances in the genetics and pathology of Lewy body dementia. Lancet Neurol. 2025;24(12):1026–1037. doi:10.1016/S1474-4422(25)00363-1

3. Ryman SG, Yutsis M, Tian L, et al. Cognition at each stage of Lewy body disease with co-occurring Alzheimer’s disease pathology. J Alzheimers Dis. 2021;80(3):1243–1256. doi:10.3233/JAD-201187

4. Abdelnour C, van Steenoven I, Londos E, et al. Alzheimer’s disease cerebrospinal fluid biomarkers predict cognitive decline in lewy body dementia. Mov Disord. 2016;31(8):1203–1208. doi:10.1002/mds.26668

5. Robinson JL, Lee EB, Xie SX, et al. Neurodegenerative disease concomitant proteinopathies are prevalent, age-related and APOE4-associated. Brain. 2018;141(7):2181–2193. doi:10.1093/brain/awy146

6. Coughlin DG, Hurtig HI, Irwin DJ. Pathological influences on clinical heterogeneity in Lewy body diseases. Mov Disord. 2020;35(1):5–19. doi:10.1002/mds.27867

7. Abdelnour C, Ferreira D, Oppedal K, et al. The combined effect of amyloid-β and tau biomarkers on brain atrophy in dementia with Lewy bodies. NeuroImage Clin. 2020;27(102333):102333. doi:10.1016/j.nicl.2020.102333

8. Blanc F, Mahmoudi R, Jonveaux T, et al. Long-term cognitive outcome of Alzheimer’s disease and dementia with Lewy bodies: dual disease is worse. Alzheimers Res Ther. 2017;9(1). doi:10.1186/s13195-017-0272-8

9. Tan JH, Laurell AA, Sidhom E, Rowe JB, O’Brien JT. The effect of Amyloid and Tau Co-pathology on disease progression in Lewy body dementia: A systematic review. Parkinsonism Relat Disord. 2025;131(107255):107255. doi:10.1016/j.parkreldis.2024.107255

10. Lemstra AW, de Beer MH, Teunissen CE, et al. Concomitant AD pathology affects clinical manifestation and survival in dementia with Lewy bodies. J Neurol Neurosurg Psychiatry. 2017;88(2):113–118. doi:10.1136/jnnp-2016-313775

11. Graff-Radford J, Boeve BF, Pedraza O, et al. Imaging and acetylcholinesterase inhibitor response in dementia with Lewy bodies. Brain. 2012;135(Pt 8):2470–2477. doi:10.1093/brain/aws173

12. Kang JH, Korecka M, Lee EB, et al. Alzheimer disease biomarkers: Moving from CSF to plasma for reliable detection of amyloid and tau pathology. Clin Chem. 2023;69(11):1247–1259. doi:10.1093/clinchem/hvad139

13. Ashton NJ, Brum WS, Di Molfetta G, et al. Diagnostic accuracy of a plasma phosphorylated tau 217 immunoassay for Alzheimer disease pathology. JAMA Neurol. 2024;81(3):255–263. doi:10.1001/jamaneurol.2023.5319

14. Palmqvist S, Warmenhoven N, Anastasi F, et al. Plasma phospho-tau217 for Alzheimer’s disease diagnosis in primary and secondary care using a fully automated platform. Nat Med. 2025;31(6):2036–2043. doi:10.1038/s41591-025-03622-w

15. Hall S, Janelidze S, Londos E, et al. Plasma phospho-tau identifies Alzheimer’s co-pathology in patients with Lewy body disease. Mov Disord. 2021;36(3):767–771. doi:10.1002/mds.28370

16. Rousset RZ, Claessen T, van Harten AC, et al. Performance of plasma p-tau217 and NfL in an unselected memory clinic setting. Alzheimers Dement (Amst). 2024;16(4):e70003. doi:10.1002/dad2.70003

17. Smith AM, Lorkiewicz SA, Arslan B, et al. Plasma phosphorylated tau 217 detects amyloid-β in neuronal synuclein disease. NPJ Parkinsons Dis. Published online April 10, 2026. doi:10.1038/s41531-026-01341-8

18. Woo KA, Yoon EJ, Kim R, et al. Plasma p-tau217 and Aβ42/40 as markers of Aβ pathology in the Lewy body continuum. Alzheimers Dement. 2025;21(10):e70788. doi:10.1002/alz.70788

19. Myers PS, O’Donnell JL, Jackson JJ, et al. Proteinopathy and longitudinal cognitive decline in Parkinson disease. Neurology. 2022;99(1):e66–e76. doi:10.1212/WNL.0000000000200344

20. Musso G, Fiorenzato E, Misenti V, et al. Detecting amyloid and tau pathology in Parkinson’s disease, 4R-tauopathies and control subjects with plasma pTau217. Front Neurol. 2025;16:1638852. doi:10.3389/fneur.2025.1638852

21. Yan S, Sahoo A, Zerenner T, et al. Serum p-tau217 is a prognostic indicator of cognitive impairment in idiopathic REM sleep behavior disorder. Ann Neurol. 2025;(ana.78109). doi:10.1002/ana.78109

22. Tropea TF, Stevenson PA, Flitter M, et al. Association between plasma P-tau217 and Alzheimer’s copathology and cognitive decline in Parkinson’s disease. Ann Neurol. 2026;(ana.78201). doi:10.1002/ana.78201

23. Shahid M, Rawls A, Ramirez V, et al. Illusory responses across the Lewy body disease spectrum. Ann Neurol. 2023;93(4):702–714. doi:10.1002/ana.26574

24. McKhann GM, Knopman DS, Chertkow H, et al. The diagnosis of dementia due to Alzheimer’s disease: Recommendations from the National Institute on Aging-Alzheimer’s Association workgroups on diagnostic guidelines for Alzheimer’s disease. Alzheimers Dement. 2011;7(3):263–269. doi:10.1016/j.jalz.2011.03.005

25. Jack CR Jr, Andrews JS, Beach TG, et al. Revised criteria for diagnosis and staging of Alzheimer’s disease: Alzheimer’s Association Workgroup. Alzheimers Dement. 2024;20(8):5143–5169. doi:10.1002/alz.13859

26. Gibb WR, Lees AJ. The relevance of the Lewy body to the pathogenesis of idiopathic Parkinson’s disease. J Neurol Neurosurg Psychiatry. 1988;51(6):745–752. doi:10.1136/jnnp.51.6.745

27. Litvan I, Goldman JG, Tröster AI, et al. Diagnostic criteria for mild cognitive impairment in Parkinson’s disease: Movement Disorder Society Task Force guidelines. Mov Disord. 2012;27(3):349–356. doi:10.1002/mds.24893

28. McKeith IG, Ferman TJ, Thomas AJ, et al. Research criteria for the diagnosis of prodromal dementia with Lewy bodies. Neurology. 2020;94(17):743–755. doi:10.1212/wnl.0000000000009323

29. Emre M, Aarsland D, Brown R, et al. Clinical diagnostic criteria for dementia associated with Parkinson’s disease. Mov Disord. 2007;22(12):1689–1707; quiz 1837. doi:10.1002/mds.21507

30. McKeith IG, Boeve BF, Dickson DW, et al. Diagnosis and management of dementia with Lewy bodies. Neurology. 2017;89(1):88–100. doi:10.1212/wnl.0000000000004058

31. Montoliu-Gaya L, Valeriano-Lorenzo E, Ashton NJ, et al. Plasma tau biomarkers are distinctly associated with tau tangles and decreased with Lewy body pathology. Alzheimers Dement. 2025;21(8):e70562. doi:10.1002/alz.70562

32. Morris JC. Clinical dementia rating: a reliable and valid diagnostic and staging measure for dementia of the Alzheimer type. Int Psychogeriatr. 1997;9 Suppl 1:173–176; discussion 177-8. doi:10.1017/s1041610297004870

33. Nasreddine ZS, Phillips NA, Bédirian V, et al. The Montreal Cognitive Assessment, MoCA: a brief screening tool for mild cognitive impairment. J Am Geriatr Soc. 2005;53(4):695–699. doi:10.1111/j.1532-5415.2005.53221.x

34. Cedarbaum JM, Jaros M, Hernandez C, et al. Rationale for use of the Clinical Dementia Rating Sum of Boxes as a primary outcome measure for Alzheimer’s disease clinical trials. Alzheimers Dement. 2013;9(1 Suppl):S45–55. doi:10.1016/j.jalz.2011.11.002

35. Weintraub S, Besser L, Dodge HH, et al. Version 3 of the Alzheimer Disease Centers’ neuropsychological test battery in the Uniform Data Set (UDS). Alzheimer Dis Assoc Disord. 2018;32(1):10–17. doi:10.1097/WAD.0000000000000223

36. Kiselica AM, Webber TA, Benge JF. The Uniform Dataset 3.0 neuropsychological battery: Factor structure, invariance testing, and demographically adjusted factor score calculation. J Int Neuropsychol Soc. 2020;26(6):576–586. doi:10.1017/S135561772000003X

37. Hayden KM, Jones RN, Zimmer C, et al. Factor structure of the National Alzheimer’s Coordinating Centers uniform dataset neuropsychological battery: an evaluation of invariance between and within groups over time. Alzheimer Dis Assoc Disord. 2011;25(2):128–137. doi:10.1097/WAD.0b013e3181ffa76d

38. Young CB, Cholerton B, Smith AM, et al. The Parkinson’s Disease Composite of Executive Functioning: A measure for detecting cognitive decline in clinical trials. Neurology. 2024;103(2):e209609. doi:10.1212/WNL.0000000000209609

39. Abdelnour C, Young CB, Shahid-Besanti M, et al. Plasma pTau181 reveals a pathological signature that predicts cognitive outcomes in Lewy body disease. Ann Neurol. 2024;96(3):526–538. doi:10.1002/ana.27003

40. The R project for statistical computing. Accessed August 27, 2025. https://www.R-project.org/

41. Donohue MC, Langford O, Insel PS, et al. Natural cubic splines for the analysis of Alzheimer’s clinical trials. Pharm Stat. 2023;22(3):508–519. doi:10.1002/pst.2285

42. Bolsewig K, van Unnik AAJM, Blujdea ER, et al. Association of plasma amyloid, P-tau, GFAP, and NfL with CSF, clinical, and cognitive features in patients with Dementia With Lewy Bodies. Neurology. 2024;102(12):e209418. doi:10.1212/WNL.0000000000209418

43. Pilotto A, Imarisio A, Carrarini C, et al. Plasma neurofilament light chain predicts cognitive progression in prodromal and clinical dementia with Lewy bodies. J Alzheimers Dis. 2021;82(3):913–919. doi:10.3233/JAD-210342

44. Donaghy PC, Firbank M, Petrides G, et al. The relationship between plasma biomarkers and amyloid PET in dementia with Lewy bodies. Parkinsonism Relat Disord. 2022;101:111–116. doi:10.1016/j.parkreldis.2022.07.008

45. Guillén N, Esteller D, Sarto J, et al. Plasma p-Tau217 and GFAP predict widespread cognitive decline in Alzheimer’s disease. J Neurol. 2025;273(1):28. doi:10.1007/s00415-025-13556-5

46. Wu CY, Chen L, Fatima H, et al. Combined use of plasma p-tau217, NfL, and GFAP predicts domain-specific cognitive decline in cognitively unimpaired and MCI individuals. Alzheimers Dement. 2025;21(12):e70934. doi:10.1002/alz.70934

47. Mattsson-Carlgren N, Janelidze S, Bateman RJ, et al. Soluble P-tau217 reflects amyloid and tau pathology and mediates the association of amyloid with tau. EMBO Mol Med. 2021;13(6):e14022. doi:10.15252/emmm.202114022

48. Janelidze S, Berron D, Smith R, et al. Associations of plasma phospho-Tau217 levels with tau positron emission tomography in early Alzheimer disease. JAMA Neurol. 2021;78(2):149–156. doi:10.1001/jamaneurol.2020.4201

49. Hamilton JM, Salmon DP, Galasko D, et al. Visuospatial deficits predict rate of cognitive decline in autopsy-verified dementia with Lewy bodies. Neuropsychology. 2008;22(6):729–737. doi:10.1037/a0012949

50. Gu Y, Kociolek A, Fernandez KK, et al. Clinical trajectories at the end of life in autopsy-confirmed dementia patients with Alzheimer disease and Lewy bodies pathologies. Neurology. 2022;98(21):e2140–e2149. doi:10.1212/WNL.0000000000200259

51. Thomas AJ, Hamilton CA, Heslegrave A, et al. A longitudinal study of plasma pTau181 in mild cognitive impairment with Lewy bodies and Alzheimer’s disease. Mov Disord. 2022;37(7):1495–1504. doi:10.1002/mds.28994

