## Supplemental materials including: eMethods, eResults, eReferences, eTables, and eFigures for "Plasma pTau217 as a Prognostic, Risk-Stratification, and Monitoring Biomarker of Clinical Progression in Lewy Body Disease"

**eMaterials**

### 1. eMethods

#### 1.1. ADRC Enrollment

All participants were enrolled in the Iqbal Farrukh and Asad Jamal Stanford Alzheimer's Disease Research Center (ADRC) study. This longitudinal, observational cohort recruited adults with Alzheimer's disease (AD) and related neurodegenerative disorders, including Parkinson's disease (PD) and dementia with Lewy bodies (DLB), as well as cognitively healthy older adults via clinician referral, community outreach, and other Stanford University longitudinal aging studies. Individuals with large vessel stroke by history or magnetic resonance imaging, toxic-metabolic encephalopathy, and cognitive impairment of psychiatric or unknown etiology were excluded.

Healthy control (HC) participants were community-dwelling older adults (ages ≥ 70 years or 40-69 years with close family history of AD and related disorders) without history of neurologic disease or cognitive impairment. Participants were required to be in reasonably good health and willing to undergo longitudinal follow-up evaluations. The presence of neurodegenerative disease and HC status was confirmed via clinical consensus (see eMethods 1.2) following enrollment.

#### 1.2. Clinical Diagnosis Groups

For participants in the current study, clinical diagnoses were established during multidisciplinary Stanford ADRC consensus meetings where at least two neurologists, one neuropsychologist, and other study personnel reviewed participant clinical data collected at each annual ADRC study visit. Cognitive status (normal cognition, mild cognitive impairment [MCI], or dementia) and etiology of cognitive impairment were determined following each participant's baseline and annual follow-up study visits. Clinical consensus procedures were blind to plasma pTau217 analysis. However, amyloid and tau PET data were available at the time of consensus diagnosis for a subgroup of participants.

Clinical diagnosis groups were defined by baseline ADRC consensus as follows:

- HC: Neurologically healthy older adults who performed within normal age- and sex-adjusted ranges on comprehensive neuropsychological testing.
- AD spectrum: Individuals with MCI or dementia due to AD according to NIH AD Diagnostic Guidelines.[1,2]
- Lewy body disease (LBD) spectrum: Individuals diagnosed with PD according to UK Brain Bank criteria[3] without objective neuropsychological test impairment, PD with MCI,[4] LBD with MCI (i.e., prodromal dementia with Lewy bodies),[5] PD dementia,[6] or dementia with Lewy bodies[7] diagnosed according to published guidelines.

#### 1.3. Plasma Collection and pTau217 Measurement

Whole blood was collected from participants annually at each Stanford ADRC study visit by venipuncture into ethylenediaminetetraacetic acid (EDTA) tubes and centrifuged (2000 x g, 10 min, 4°C). In our sample, baseline plasma was collected a median of 8 days (IQR: 0-53 days) from consensus diagnosis, and within 6 months of consensus diagnosis in 97% of participants. Isolated plasma was then aliquoted and stored at -80°C for future biomarker analysis.

Samples were not thawed prior to the pTau217 assay. For the pTau217 assay, plasma samples were thawed at room temperature (RT), vortexed (2000 rpm, 30 sec., RT), and centrifuged (4000 x g, 10 min., RT). pTau217 protein levels were identified using the Quanterix Simoa® HD-X platform and ALZpath Simoa® pTau217 V2 assay kit (#104371, Quanterix). The ALZpath Simoa® pTau217 V2 assay was completed at the Department of Psychiatry and Neurochemistry at the University of Gothenburg. Assay workflow was followed as manufacturer instructed and previously published.[8]

Briefly, the ALZpath Simoa® pTau217 assay uses a proprietary monoclonal antibody specific to pTau217, paired with an N-terminal detection antibody and peptide calibrators. The calibrators were run in duplicate, with replicates masked if outside of the precision limit before being fit to a standard curve. Plasma samples were diluted three-fold and results were adjusted for dilution. Three internal quality-control levels were tested in duplicates at the beginning and end of each run. Samples outside the quantifiable range for the limit of detection (0.0072-30.0 pg/mL) were excluded from analyses.

**1.4. Plasma pTau217 Cut-Point Development**

Development of the cut-point used to define abnormal plasma pTau217 (0.54pg/mL) is described in detail in Smith et al., (2026)[9]. In brief, while plasma biomarker diagnostic cut-points are well-described in clinical AD, cut-points can vary between clinical populations of interest, particularly those with underlying Lewy body pathology. Therefore, diagnostic cut-points for detecting amyloid-β (Aβ) positivity in participants with biologically defined Neuronal Synuclein Disease (NSD)[10,11] were examined across several plasma biomarkers.

Participants were classified as NSD positive (NSD+) or negative (NSD-) using dichotomous results from the cerebrospinal fluid (CSF) α-synuclein seed amplification (SAA) assay (SYNTap® Biomarker Test) and as amyloid positive (Aβ+) or negative (Aβ-) using the CSF Aβ 42/40 ratio (Lumipulse G1200 platform). The CSF Aβ 42/40 ratio cut-point was calculated using amyloid PET as the reference standard. That is, in an independent subset of Stanford cohort participants with amyloid PET data, Gaussian mixture modeling identified a positivity threshold of 36 centiloids. Based on this 36 centiloid amyloid PET threshold, ROC curve analyses and the Youden index identified a CSF Aβ 42/40 positivity cut-point of < 0.09.

This yielded four groups across which plasma biomarker performance was tested: NSD+/Aβ+ ($n$ = 28), NSD+/ Aβ- ($n$ = 39), NSD-/ Aβ+ ($n$ = 44), and NSD-/ Aβ- ($n$ = 69). Separate ROC curve analyses identified plasma pTau217 as independently demonstrating the best overall diagnostic performance for detecting Aβ in NSD+ participants (AUC = 0.92, 95% CI = [0.85, 1.00]) and the largest median fold-change between Aβ+ and Aβ- participants (2.79 [SD = 0.33], 95% CI = [2.3, 3.61]). Inclusion of other biomarkers did not improve the performance of plasma pTau217.

Among NSD+ participants, separate ROC curve analyses with the Youden index identified a one-reference plasma pTau217 cut-point of > 0.54pg/mL as differentiating between Aβ+ and Aβ- participants and indicative of Aβ positivity.

In the present study, we therefore defined raw pTau217 values > 0.54pg/mL as abnormal (pTau217+) and raw values ≤ 0.54pg/mL as normal (pTau217-).

##### 1.5. Longitudinal Clinical Outcomes

###### 1.5.1. Clinical Dementia Rating Scale Sum of Boxes

The Clinical Dementia Rating-Sum of Boxes (CDR-SB) is derived from clinician ratings in six domains: memory, orientation, judgement/problem solving, community affairs, home/ hobbies, and personal care. Using semi-structured interviews with the participant and an informant,[12] each domain is rated on a 0 to 3 scale and summed to yield a continuous CDR-SB score ranging from 0 to 18 (i.e., higher scores indicate greater impairment).[13]

The CDR-SB is a widely used clinical outcome capturing functional disability and cognitive decline, and is sensitive to clinical progression in early disease stages.[14] While CDR-SB scores are comprised of both cognitive and functional features, we examine CDR-SB as a proxy of daily functioning and included other nuanced indices of cognition.

###### 1.5.2. Montreal Cognitive Assessment

The Montreal Cognitive Assessment (MoCA) is a brief screening measure of global cognition sampling multiple cognitive domains including attention, executive function, memory, language, visuospatial skills, and orientation. Each sub-test is scored and added together to yield a continuous MoCA score ranging from 0 to 30 (i.e., higher scores indicate better cognition).[15] The MoCA has shown to be a sensitive indicator of global cognitive status in LBD cohorts.[16]

###### 1.5.3. Domain-Specific Cognitive Indices

Five cognitive indices were generated by averaging *z*-scores from tests measuring abilities in each domain as follows:

a.) Memory index: Hopkins Verbal Learning Test-Revised delayed recall,[17] Craft Story 21 delayed recall,[18] Benson Complex Figure delayed recall,[19] and Free and Cued Selective Reminding Test delayed free recall.[20] Three out of four test scores were required to generate this index.

b.) Executive function index: Number Span Forward total score,[21] Number Span Backward total score, Letter-Number Sequencing total score,[22] Victoria Stroop color-word trial total time score,[23] Trail Making Task B total time score,[24] and Phonemic Fluency total score.[25–28] Four out of six scores were required to generate this index.

c.) Processing speed index: Trail Making Test A total time score, Victoria Stroop color-naming trial total time score, Victoria Stroop word-reading trial total time score, and Coding total score.[22] Three out of four scores were required for this index.

d.) Language index: Semantic Fluency total score,[29] Multilingual Naming Test (MINT) total score,[30] and Cookie Theft total score.[31,32] Two out of three scores were required for this index.

e.) Visuospatial function index: Judgement of Line Orientation Task total score,[33] Benson Complex Figure copy trial, Clock Drawing Test copy trial,[34] and the Clock Drawing Test command trial. Two out of four scores were required for this index.

#### 1.6. Statistical Analyses

##### 1.6.1. Plasma pTau217 as a Prognostic and Monitoring Biomarker

The following model was fit for clinical outcomes where longitudinal change was best modeled linearly (CDR-SB, MoCA, memory index, language index) or using a linear time specification:

$$
\begin{aligned}
(1)\ Y_{it} = {} & \beta_0 + \beta_1(\text{time}_{it}) + \beta_2(\text{age}_i) + \beta_3(\text{sex}_i) + \beta_4(\text{education}_i) + \beta_5(\text{diagnosis group}_i) \\
& + \beta_6(\text{baseline pTau217}_i) + \beta_7(\text{pTau217 slope}_i) \\
& + \beta_8(\text{baseline pTau217}_i \cdot \text{time}_{it}) + \beta_9(\text{pTau217 slope}_i \cdot \text{time}_{it}) + \beta_{10}(\text{diagnosis group}_i \cdot \text{time}_{it}) \\
& + \beta_{11}(\text{baseline pTau217}_i \cdot \text{diagnosis group}_i) + \beta_{12}(\text{pTau217 slope}_i \cdot \text{diagnosis group}_i) \\
& + \beta_{13}(\text{baseline pTau217}_i \cdot \text{diagnosis group}_i \cdot \text{time}_{it}) \\
& + \beta_{14}(\text{pTau217 slope}_i \cdot \text{diagnosis group}_i \cdot \text{time}_{it}) \\
& + b_{0i} + b_{1i}\text{time}_{it} + \varepsilon_{it}
\end{aligned}
$$

Where *i* represents an individual participant and *t* represents years since initial assessment, βs are estimates for fixed effects, $Y_{it}$ is the observed clinical outcome, $\varepsilon_{it}$ is residual error, and random effects are represented by $b_{0i}$ (intercept) and $b_{1i}\text{time}_{it}$ (slope). Primary predictors of interest were baseline $\text{pTau217}_i$ ($\beta_6$) and pTau217 $\text{slope}_i$ ($\beta_7$). All models included interactions between pTau217 predictors, clinical diagnosis group ($\beta_5$), and time ($\beta_{13\text{-}14}$). Two-way interactions were retained in each model ($\beta_{8\text{-}}\ \beta_{12}$). Covariates included age, sex, and education ($\beta_{2\text{-}4}$)

For clinical outcomes demonstrating non-linear longitudinal change (executive function index, visuospatial function index, processing speed index), time was modeled using natural cubic splines with three basis functions (i.e., $S_1$, $S_2$, and $S_3$). Spline basis functions were included as main effect and interaction terms, with all other model parameters analogous to the linear specification (1). The following model was fit:

$$
\begin{aligned}
(2)\ Y_{it} = {} & \beta_0 + \beta_1 S_1(\text{time}_{it}) + \beta_2 S_2(\text{time}_{it}) + \beta_3 S_3(\text{time}_{it}) + \beta_4(\text{age}_i) + \beta_5(\text{sex}_i) + \beta_6(\text{education}_i) + \beta_7(\text{diagnosis group}_i) \\
& + \beta_8(\text{baseline pTau217}_i) + \beta_9(\text{pTau217 slope}_i) \\
& + \beta_{10}(\text{diagnosis group}_i \cdot S_1(\text{time}_{it})) + \beta_{11}(\text{diagnosis groupi} \cdot S_2(\text{time}_{it})) + \beta_{12}(\text{diagnosis group}_i \cdot S_3(\text{time}_{it})) \\
& + \beta_{13}(\text{baseline pTau217}_i \cdot S_1(\text{time}_{it})) + \beta_{14}(\text{baseline pTau217}_i \cdot S_2(\text{time}_{it})) + \beta_{15}(\text{baseline pTau217}_i \cdot \\
& S_3(\text{time}_{it})) \\
& + \beta_{16}(\text{pTau217 slope}_i \cdot S_1(\text{time}_{it})) + \beta_{17}(\text{pTau217 slope}_i \cdot S_2(\text{time}_{it})) + \beta_{18}(\text{pTau217 slope}_i \cdot S_3(\text{time}_{it})) \\
& + \beta_{19}(\text{baseline pTau217}_i \cdot \text{diagnosis group}_i) + \beta_{20}(\text{pTau217 slope}_i \cdot \text{diagnosis group}_i) \\
& + \beta_{21}(\text{baseline pTau217}_i \cdot \text{diagnosis group}_i \cdot S_1(\text{time}_{it})) + \beta_{22}(\text{baseline pTau217}_i \cdot \text{diagnosis group}_i \cdot \\
& S_2(\text{time}_{it})) + \beta_{23}(\text{baseline pTau217}_i \cdot \text{diagnosis group}_i \cdot S_3(\text{time}_{it}))
\end{aligned}
$$

$$+ \beta_{24}(\text{pTau217 slope}_i \cdot \text{diagnosis group}_i \cdot S_1(\text{time}_{it})) + \beta_{25}(\text{pTau217 slope}_i \cdot \text{diagnosis group}_i \cdot S_2(\text{time}_{it}))$$
$$+ \beta_{26}(\text{pTau217 slope}_i \cdot \text{diagnosis group}_i \cdot S_3(\text{time}_{it}))$$
$$+ b_{0i} + b_{1i}\text{time}_{it} + \varepsilon_{it}$$

##### 1.6.2. Sensitivity Analyses of Non-Linear Baseline pTau217 Effects

Non-linear associations between plasma pTau217 and longitudinal clinical outcome change were evaluated for all seven clinical outcomes. Specifically, two non-linear baseline pTau217 parameterizations were included separately in the linear mixed-effects models described in section 1.6.1 while otherwise retaining the original model structure. Linear baseline pTau217 and pTau217 slope terms were retained in sensitivity models to enable comparisons with corresponding linear baseline pTau217 models. Quadratic baseline pTau217 terms tested whether associations with longitudinal clinical outcome change varied in a curvilinear manner across the baseline pTau217 distribution. In separate models, piecewise baseline pTau217 terms with knots at the 25th, 50th, and 75th percentiles were included to test whether associations with longitudinal clinical outcome change differed across lower, middle, or higher ranges of baseline pTau217 without imposing a smooth quadratic function.

Bayesian Information Criterion (BIC) and likelihood ratio tests evaluated whether inclusion of non-linear baseline pTau217 terms improved model fit compared to corresponding linear baseline pTau217 models. Additional sensitivity analyses examined whether inclusion of non-linear baseline pTau217 terms changed interpretation of primary findings. That is, interactions between linear baseline pTau217 or pTau217 slope, clinical diagnosis group, and time evaluated effects of pTau217 on longitudinal clinical outcome change. We focused on our primary disease group of interest, LBD, by deriving LBD-specific simple slope estimates for linear baseline pTau217 and pTau217 slope effects using Wald tests with Benjamini-Hochberg false discovery rate corrections. Direction, magnitude, and statistical significance of estimates were qualitatively compared to those from original linear baseline pTau217 models.

##### 1.6.3. Plasma pTau217 as a Risk-Stratification Biomarker of Faster Cognitive and Functional Decline

Complementary linear mixed-effects models were specified using baseline pTau217 status (abnormal versus normal) instead of baseline pTau217 and pTau217 slope as the primary predictor of interest. For clinical outcomes best characterized by linear longitudinal change (CDR-SB, MoCA, memory index, and language index) the following model was fit:

$$(3)\ Y_{it} = \beta_0 + \beta_1(\text{time}_{it}) + \beta_2(\text{age}_i) + \beta_3(\text{sex}_i) + \beta_4(\text{education}_i) + \beta_5(\text{diagnosis group}_i)$$
$$+ \beta_6(\text{baseline pTau217 Status}_i) +$$
$$+ \beta_7(\text{baseline pTau217 Status}_i \cdot \text{time}_{it}) + \beta_8(\text{diagnosis group}_i \cdot \text{time}_{it})$$
$$+ \beta_9(\text{baseline pTau217 Status}_i \cdot \text{diagnosis group}_i)$$
$$+ \beta_{10}(\text{baseline pTau217 Status}_i \cdot \text{diagnosis group}_i \cdot \text{time}_{it})$$
$$+ b_{0i} + b_{1i}\text{time}_{it} + \varepsilon_{it}$$

For clinical outcomes best characterized by non-linear change (executive function index, visuospatial function index, processing speed index), the following model was fit:

$$(4)\ Y_{it} = \beta_0 + \beta_1 S_1(\text{time}_{it}) + \beta_2 S_2(\text{time}_{it}) + \beta_3 S_3(\text{time}_{it}) + \beta_4(\text{age}_i) + \beta_5(\text{sex}_i) + \beta_6(\text{education}_i) + \beta_7(\text{diagnosis group}_i)$$

$+ \beta_8$(baseline pTau217 Status$_i$) +
$+ \beta_{10}$(diagnosis group$_i \cdot S_1$(time$_{it}$)) $+ \beta_{11}$(diagnosis group$_i \cdot S_2$(time$_{it}$)) $+ \beta_{12}$(diagnosis group $_i \cdot S_3$(time$_{it}$))
$+ \beta_{13}$(baseline pTau217 Status$_i \cdot S_1$(time$_{it}$)) $+ \beta_{14}$(baseline pTau217 Status$_i \cdot S_2$(time$_{it}$)) +
$\beta_{15}$(baseline pTau217 Status$_i \cdot S_3$(time$_{it}$))
$+ \beta_{16}$(baseline pTau217 Status$_i$ · diagnosis group$_i$)
$+ \beta_{17}$(baseline pTau217 Status$_i$ · diagnosis group$_i \cdot S_1$(time$_{it}$)) $+ \beta_{18}$(baseline pTau217 Status$_i$ ·
Diagnosis group$_i \cdot S_2$(time$_{it}$)) $+ \beta_{19}$(baseline pTau217 Status$_i$ · diagnosis group$_i \cdot S_3$(time$_{it}$))
$+ b_{0i} + b_{1i}\text{time}_{it} + \varepsilon_{it}$

Interactions between baseline pTau217 status, clinical diagnosis group, and time evaluated effects of pTau217 status on clinical outcome trajectories in each clinical diagnosis group. Using two-sided Satterthwaite *t*-tests, simple slope contrasts compared annual clinical outcome change between abnormal (> 0.54 pg/mL) and normal (≤ 0.54 pg/mL) pTau217 status groups for each clinical diagnosis group, with FDR corrections applied across these three planned contrasts. For spline models, estimates were calculated by first generating model-estimated total change from baseline up to 8 years, then dividing total change by 8 to calculate average annualized change.

##### 1.6.4. Sensitivity Analyses Stratifying Cox Proportional Hazard Models by Baseline Cognitive Status

Given known associations with disease progression,[35] baseline cognitive status was included in sensitivity analyses. Specifically, Cox models were stratified by baseline cognitive status using the R Survival package strata() function[36] which allowed the baseline hazard to vary across strata while preserving estimation of clinical diagnosis by pTau217 status group effects. Results were similar to those from non-stratified models and described in eResults 2.3 and eTable 8. Since baseline cognitive status is closely aligned with the conversion event, non-stratified models were retained to prevent over-correction as well as to preserve parsimony and interpretability.

#### 2. eResults

##### 2.1. Clinical Outcome Characteristics

Baseline data were available for 431-477 participants depending on clinical outcome, with variation in sample size for each outcome related to attrition, differences in participant ability to complete cognitive and functional tasks (e.g., precluded by severity of cognitive impairment), changes in ADRC study recruitment and/or protocol (e.g., COVID-19 pandemic-related changes), and inconsistent inclusion of supplemental tasks in the Stanford ADRC neuropsychological test battery. Sample sizes for each clinical outcome stratified by clinical diagnosis group are shown in eTable 2.

Mean (SD) number of follow-up assessments across ADRC study visits for each clinical outcome were as follows: CDR-SB = 5.7 (2.6), MoCA = 2.9 (1.4), memory index = 3.3 (1.5), executive function index = 3.2 (1.5), visuospatial function index = 3.3 (1.6), language index = 3.5 (1.6), and processing speed index = 2.9 (1.6).

##### 2.2. Sensitivity Analyses of Non-Linear Baseline pTau217 Effects

While inclusion of quadratic and piecewise baseline pTau217 terms resulted in higher log-likelihoods and statistically significant likelihood-ratio tests, BIC values also increased, most notably after including piecewise baseline pTau217 (eTable 5). These findings suggested improvements in fit did not consistently justify use of more complex non-linear models.

Nevertheless, to examine whether inclusion of non-linear baseline pTau217 terms changed interpretation of primary findings, follow-up sensitivity analyses were conducted using models with quadratic baseline pTau217 as they were more parsimonious and interpretable. Sensitivity analyses found that higher linear baseline pTau217 was associated with faster increase in CDR-SB ($\beta = 0.38$, $p < 0.001$) and faster declines in MoCA ($\beta = -0.47$, $p < 0.001$), memory ($\beta = -0.10$, $p = 0.001$), executive function ($\beta = -2.26$, $p < 0.001$), and visuospatial function ($\beta = -4.19$, $p < 0.001$). Steeper pTau217 slope was also associated with faster increase in CDR-SB ($\beta = 0.20$, $p = 0.025$). Compared to findings from original models, one isolated change was noted where higher linear baseline pTau217 was more strongly associated with faster processing speed decline in the quadratic baseline pTau217 model ($\beta = -1.92$, $p = 0.012$; eTable 6).

##### 2.3. Sensitivity Analyses Stratifying Cox Proportional Hazard Models by Baseline Cognitive Status

In age and sex-adjusted Cox models stratified by baseline cognitive status (cognitively unimpaired versus impaired [MCI]; $n = 371$; 80 events), all proportional hazards assumptions were met. All groups had significantly higher conversion risk compared to HC pTau217- (eTable 8). Pairwise contrasts found LBD pTau217+ had a three-fold higher conversion risk compared to LBD pTau217- (HR = 3.8; 95% CI = [1.7, 8.3]; $p = 0.001$).

#### 3. References


1. McKhann GM, Knopman DS, Chertkow H, et al. The diagnosis of dementia due to Alzheimer's disease: Recommendations from the National Institute on Aging-Alzheimer's Association workgroups on diagnostic guidelines for Alzheimer's disease. *Alzheimers Dement*. 2011;7(3):263-269.

2. Jack CR Jr, Andrews JS, Beach TG, et al. Revised criteria for diagnosis and staging of Alzheimer's disease: Alzheimer's Association Workgroup. *Alzheimers Dement*. 2024;20(8):5143-5169.

3. Gibb WR, Lees AJ. The relevance of the Lewy body to the pathogenesis of idiopathic Parkinson's disease. *J Neurol Neurosurg Psychiatry*. 1988;51(6):745-752.

4. Litvan I, Goldman JG, Tröster AI, et al. Diagnostic criteria for mild cognitive impairment in Parkinson's disease: Movement Disorder Society Task Force guidelines. *Mov Disord*. 2012;27(3):349-356.

5. McKeith IG, Ferman TJ, Thomas AJ, et al. Research criteria for the diagnosis of prodromal dementia with Lewy bodies. *Neurology*. 2020;94(17):743-755.

6. Emre M, Aarsland D, Brown R, et al. Clinical diagnostic criteria for dementia associated with Parkinson's disease. *Mov Disord*. 2007;22(12):1689-1707; quiz 1837.

7. McKeith IG, Boeve BF, Dickson DW, et al. Diagnosis and management of dementia with Lewy bodies. *Neurology*. 2017;89(1):88-100.

8. Ashton NJ, Brum WS, Di Molfetta G, et al. Diagnostic accuracy of a plasma phosphorylated tau 217 immunoassay for Alzheimer disease pathology. *JAMA Neurol*. 2024;81(3):255-263.

9. Smith AM, Lorkiewicz SA, Arslan B, et al. Plasma phosphorylated tau 217 detects amyloid-β in neuronal synuclein disease. *NPJ Parkinsons Dis*. Published online April 10, 2026. doi:10.1038/s41531-026-01341-8

10. Simuni T, Chahine LM, Poston K, et al. A biological definition of neuronal α-synuclein disease: towards an integrated staging system for research. *Lancet Neurol*. 2024;23(2):178-190.

11. Simuni T, Gochanour C, Nair AR, et al. Neuronal α-synuclein disease stage progression over 5 years. *Mov Disord*. 2025;40(7):1318-1330.

12. Morris JC. Clinical dementia rating: a reliable and valid diagnostic and staging measure for dementia of the Alzheimer type. *Int Psychogeriatr*. 1997;9 Suppl 1:173-176; discussion 177-8.

13. O'Bryant SE, Waring SC, Cullum CM, et al. Staging dementia using Clinical Dementia Rating Scale Sum of Boxes scores: a Texas Alzheimer's research consortium study. *Arch Neurol*. 2008;65(8):1091-1095.

14. Cedarbaum JM, Jaros M, Hernandez C, et al. Rationale for use of the Clinical Dementia Rating Sum of Boxes as a primary outcome measure for Alzheimer's disease clinical trials. *Alzheimers Dement*. 2013;9(1 Suppl):S45-55.

15. Nasreddine ZS, Phillips NA, Bédirian V, et al. The Montreal Cognitive Assessment, MoCA: a brief screening tool for mild cognitive impairment. *J Am Geriatr Soc*. 2005;53(4):695-699.

16. Biundo R, Weis L, Bostantjopoulou S, et al. MMSE and MoCA in Parkinson's disease and dementia with Lewy bodies: a multicenter 1-year follow-up study. *J Neural Transm (Vienna)*. 2016;123(4):431-438.

17. Benedict RHB, Schretlen D, Groninger L, Brandt J. Hopkins verbal learning test – revised: Normative data and analysis of inter-form and test-retest reliability. *Clin Neuropsychol*. 1998;12(1):43-55.

18. Craft S, Newcomer J, Kanne S, et al. Memory improvement following induced hyperinsulinemia in Alzheimer's disease. *Neurobiol Aging*. 1996;17(1):123-130.

19. Possin KL, Laluz VR, Alcantar OZ, Miller BL, Kramer JH. Distinct neuroanatomical substrates and cognitive mechanisms of figure copy performance in Alzheimer's disease and behavioral variant frontotemporal dementia. *Neuropsychologia*. 2011;49(1):43-48.

20. Ivnik RJ, Smith GE, Lucas JA, Tangalos EG, Kokmen E, Petersen RC. Free and cued selective reminding test: Moans norms. *J Clin Exp Neuropsychol*. 1997;19(5):676-691.

21. Monsell SE, Dodge HH, Zhou XH, et al. Results from the NACC uniform data set neuropsychological battery Crosswalk Study. *Alzheimer Dis Assoc Disord*. 2016;30(2):134-139.

22. Wechsler D. *Wechsler Adult Intelligence Scale-Fourth Edition Administration Scoring Manual*. Psychological Corporation; 2008.

23. Troyer AK, Leach L, Strauss E. Aging and response inhibition: Normative data for the Victoria Stroop Test. *Neuropsychol Dev Cogn B Aging Neuropsychol Cogn*. 2006;13(1):20-35.

24. Reitan RM. Validity of the trail making test as an indicator of organic brain damage. *Percept Mot Skills*. 1958;8(3):271-276.

25. Benton, A. L., Hamsher, K. D., & Sivan, A. B. *Multilingual Aphasia Examination*.; 1976.

26. Ruff RM, Light RH, Parker SB, Levin HS. Benton controlled oral word association test: Reliability and updated norms. *Arch Clin Neuropsychol*. 1996;11(4):329-338.

27. Gustavson DE, Panizzon MS, Franz CE, et al. Integrating verbal fluency with executive functions: Evidence from twin studies in adolescence and middle age. *J Exp Psychol Gen*. 2019;148(12):2104-2119.

28. Troyer AK, Moscovitch M, Winocur G, Alexander MP, Stuss D. Clustering and switching on verbal fluency: the effects of focal frontal- and temporal-lobe lesions. *Neuropsychologia*. 1998;36(6):499-504.

29. Morris JC, Heyman A, Mohs RC, et al. The Consortium to Establish a Registry for Alzheimer's Disease (CERAD). Part I. Clinical and neuropsychological assessment of Alzheimer's disease. *Neurology*. 1989;39(9):1159-1165.

30. Ivanova I, Salmon DP, Gollan TH. The multilingual naming test in Alzheimer's disease: clues to the origin of naming impairments. *J Int Neuropsychol Soc*. 2013;19(3):272-283.

31. Giles E, Patterson K, Hodges JR. Performance on the Boston Cookie theft picture description task in patients with early dementia of the Alzheimer's type: Missing information. *Aphasiology*. 1996;10(4):395-408.

32. Goodglass H, Kaplan E. *Boston Diagnostic Aphasia Examination: Boston Naming Test*. 3rd ed. Lippincott Williams and Wilkins; 2000.

33. Benton S, Hamsher K. *Contributions Neuropsychological Assessment. Clinical Manual*. Oxford University Press; 1994.

34. Rouleau I, Salmon DP, Butters N, Kennedy C, McGuire K. Quantitative and qualitative analyses of clock drawings in Alzheimer's and Huntington's disease. *Brain Cogn*. 1992;18(1):70-87.

35. Pedersen KF, Larsen JP, Tysnes OB, Alves G. Prognosis of mild cognitive impairment in early Parkinson disease: the Norwegian ParkWest study. *JAMA Neurol*. 2013;70(5):580-586.

36. Therneau T. *A Package for Survival Analysis in R*.; 2024. https://CRAN.R-project.org/package=survival

#### 4. eTables

**eTable 1.** Descriptive data for raw and $log_{10}$-transformed plasma pTau217 over time (*N* = 501)

| Annual ADRC Study Visit Number | Sample Size Per Year | Raw pTau217 pg/mL *Median (IQR)* | pTau217 $log_{10}$ transformed *Median (IQR)* |
|---|---|---|---|
| 1 | *n* = 493 | 0.00 (0.31, 0.85) | -0.32 (-0.51, -0.07) |
| 2 | *n* = 289 | 0.00 (0.30, 0.84) | -0.32 (-0.52, -0.08) |
| 3 | *n* = 141 | 0.00 (0.34, 0.93) | -0.32 (-0.47, -0.03) |
| 4 | *n* = 45 | 0.00 (0.37, 0.81) | -0.32 (-0.43, -0.09) |
| 5 | *n* = 7 | 1.00 (0.42, 1.13) | -0.19 (-0.38, 0.04) |

**Note.** Abbreviations: ADRC = Alzheimer's Disease Research Center; IQR = Interquartile range. At ADRC study visit 1, eight participants were missing pTau217 data, and pTau217 was analyzed starting at study visit 2. Sample sizes otherwise varied between ADRC study visits 2-5 due to out-of-range plasma pTau217 measurements and plasma pTau217 not being measured at the same study visit for all participants. $Log_{10}$-transformed values are negative when raw values were < 1 pg/mL.

**eTable 2.** Sample sizes over time for clinical outcomes stratified by clinical diagnosis group

| Clinical Diagnosis Group | Clinical Outcome | ADRC Study Visit Year | | | | | | | | |
|---|---|---|---|---|---|---|---|---|---|---|
| | | BL | 1 | 2 | 3 | 4 | 5 | 6 | 7 | 8 |
| HC | CDR-SB | 221 | 218 | 193 | 154 | 135 | 120 | 84 | 51 | 21 |
| | MoCA | 219 | 226 | 139 | 16 | 80 | 5 | 17 | 6 | 2 |
| | Memory | 233 | 226 | 139 | 15 | 80 | 7 | 17 | 6 | 2 |
| | Executive Function | 233 | 226 | 139 | 14 | 78 | 6 | 17 | 6 | 2 |
| | Visuospatial Function | 229 | 217 | 142 | 15 | 87 | 7 | 31 | 5 | 2 |
| | Language | 233 | 231 | 147 | 16 | 92 | 7 | 31 | 6 | 2 |
| | Processing Speed | 223 | 214 | 126 | 13 | 61 | 7 | 14 | 5 | 2 |
| LBD | CDR-SB | 125 | 124 | 103 | 87 | 72 | 59 | 30 | 14 | 6 |
| | MoCA | 115 | 124 | 89 | 26 | 35 | 17 | 23 | 12 | 10 |
| | Memory | 126 | 123 | 88 | 26 | 34 | 15 | 22 | 12 | 10 |
| | Executive Function | 123 | 120 | 87 | 25 | 34 | 15 | 22 | 11 | 9 |
| | Visuospatial Function | 124 | 116 | 89 | 31 | 48 | 16 | 22 | 11 | 10 |
| | Language | 126 | 126 | 93 | 31 | 48 | 16 | 24 | 12 | 10 |
| | Processing Speed | 114 | 111 | 74 | 20 | 26 | 13 | 19 | 11 | 9 |
| AD | CDR-SB | 122 | 117 | 91 | 56 | 46 | 33 | 17 | 13 | 4 |
| | MoCA | 123 | 117 | 67 | 39 | 22 | 10 | 6 | 3 | 3 |
| | Memory | 116 | 109 | 60 | 36 | 20 | 8 | 5 | 3 | 3 |
| | Executive Function | 107 | 99 | 53 | 33 | 18 | 8 | 5 | 3 | 3 |
| | Visuospatial Function | 112 | 104 | 56 | 36 | 23 | 13 | 6 | 3 | 2 |
| | Language | 118 | 114 | 57 | 41 | 22 | 13 | 6 | 3 | 3 |
| | Processing Speed | 94 | 90 | 40 | 28 | 13 | 6 | 5 | 2 | 2 |

**Note.** Abbreviations: AD = Alzheimer's disease spectrum; BL = baseline; HC = Healthy controls; LBD = Lewy body disease spectrum. Values represent the number of participants with available data at each annual ADRC study visit for each clinical outcome stratified by clinical diagnosis group. Sample sizes vary across clinical outcomes and timepoints due to differences in missingness. BL values for each clinical outcome were defined as first assessment across ADRC study visits and excluded participants with missing data at all study visits.

**eTable 3.** Mean follow-up duration for longitudinal clinical outcomes by clinical diagnosis

| Clinical Outcome | HC | LBD | AD | Test Statistic | *p*-value |
|---|---|---|---|---|---|
| CDR-SB, *n* | 200 | 108 | 93 | | |
| Follow-Up[a] | 4.2 (2.0) | 4.0 (1.8) | 3.2 (1.9) | 17.8 | < 0.001 |
| MoCA, *n* | 166 | 92 | 68 | | |
| Follow-Up | 3.4 (2.1) | 4.2 (2.4) | 2.9 (1.8) | 11.2 | 0.004 |
| Cognitive Indices | | | | | |
| Memory, *n* | 165 | 91 | 59 | | |
| Follow-Up | 3.4 (2.2) | 4.1 (2.5) | 2.9 (1.8) | 7.6 | 0.022 |
| Executive Function, *n* | 165 | 90 | 52 | | |
| Follow-Up | 3.3 (2.2) | 4.1 (2.4) | 2.9 (1.8) | 7.9 | 0.020 |
| Visuospatial Function, *n* | 170 | 95 | 57 | | |
| Follow-Up | 3.4 (2.0) | 4.0 (2.3) | 2.8 (1.9) | 10.4 | 0.005 |
| Processing Speed, *n* | 154 | 82 | 47 | | |
| Follow-Up | 3.3 (2.1) | 4.1 (2.4) | 2.8 (1.8) | 9.3 | 0.010 |
| Language, *n* | 171 | 96 | 59 | | |
| Follow-Up | 3.5 (2.01) | 4.1 (2.4) | 2.9 (1.8) | 9.2 | 0.010 |
| CDR-SB, *n* | 200 | 108 | 93 | | |

**Note.** Abbreviations: *n* = sample size for participants with ≥ 2 observations across all timepoints for a given clinical outcome. Follow-up duration in years was calculated separately for each clinical outcome. Kruskal-Wallis tests compared follow-up duration across clinical diagnostic groups for each clinical outcome.

[a]Follow-up duration was defined as years between first and last available assessment and reported as mean (standard deviation) years.

**eTable 4.** Fit indices for mixed effects models including linear and natural cubic spline specifications for time

| pTau217 Predictor Variable | Clinical Outcome | AIC | | BIC | | Log Likelihood | | *p*-value |
|---|---|---|---|---|---|---|---|---|
| | | Linear | Spline | Linear | Spline | Linear | Spline | |
| pTau217 Baseline and Slope | CDR-SB[a] | 4547.0 | 4543.7 | 4578.7 | 4586.0 | 4535.0 | 4527.7 | 0.026 |
| ($N$ = 290) | MoCA[a] | 3997.3 | 3997.6 | 4025.0 | 4034.6 | 3985.3 | 3981.6 | 0.158 |
| | Cognitive Indices | | | | | | | |
| | Memory[a] | 2103.4 | 2105.0 | 2131.9 | 2143.0 | 2091.4 | 2088.9 | 0.299 |
| | Executive[b] | 1496.9 | 1480.4 | 1525.3 | 1518.2 | 1484.9 | 1464.4 | < 0.001 |
| | Visuospatial[b] | 2652.2 | 2642.2 | 2680.9 | 2680.5 | 2640.2 | 2626.2 | < 0.001 |
| | Language[a] | 2254.5 | 2258.1 | 2283.5 | 2296.8 | 2242.4 | 2242.1 | 0.852 |
| | Processing Speed[b] | 1679.1 | 1671.7 | 1706.8 | 1708.6 | 1667.1 | 1655.7 | 0.003 |
| Baseline pTau217 Status | CDR-SB[a] | 6566.2 | 6563.8 | 6599.5 | 6608.2 | 6554.2 | 6547.8 | 0.041 |
| ($N$ = 501) | MoCA[a] | 5521.9 | 5523.8 | 5551.2 | 5563.0 | 5509.9 | 5507.8 | 0.359 |
| | Cognitive Indices | | | | | | | |
| | Memory[a] | 3032.1 | 3035.9 | 3062.4 | 3076.2 | 3020.1 | 3019.9 | 0.880 |
| | Executive[b] | 2203.1 | 2182.6 | 2233.1 | 2222.7 | 2191.1 | 2166.6 | <0.001 |
| | Visuospatial[b] | 3693.2 | 3687.3 | 3723.6 | 3727.8 | 3681.2 | 3671.3 | 0.007 |
| | Language[a] | 3096.7 | 3098.7 | 3127.3 | 3139.6 | 3084.6 | 3082.7 | 0.382 |
| | Processing Speed[b] | 2550.5 | 2539.7 | 2579.9 | 2578.8 | 2538.5 | 2523.7 | <0.001 |

**Note.** Abbreviations: AIC = Akaike Information Criterion; BIC = Bayesian Information Criterion; CDR-SB = Clinical Dementia Rating-Sum of Boxes; Executive = Executive function; MoCA = Montreal Cognitive Assessment; Visuospatial = Visuospatial function. Lower AIC and BIC values indicated better fit. Nested likelihood ratio $\chi^2$ tests compared linear versus spline models, where $p < 0.05$ indicated better fit for the spline model. Models analyzing baseline pTau217 and pTau217 slope variables included participants with ≥ 2 plasma measurements whereas models analyzing baseline pTau217 status included participants with ≥ 1 plasma measurement.
[a]Final model used linear time specification, clinical outcome trajectories were approximately linear.
[b]Final model used natural cubic spline specification for time, clinical outcome trajectories were non-linear.

**eTable 5.** Fit indices comparing linear mixed-effects models with linear and non-linear baseline pTau217 parameterizations

| Clinical Outcome | Baseline pTau217 Parameterization | BIC | Log-likelihood | $\chi^2$ | df | *p*-value |
|---|---|---|---|---|---|---|
| CDR-SB | Linear[a] | 4478.3 | -2148.1 | | | |
| | Quadratic[b] | 4495.2 | -2134.6 | 26.84 | 6 | < 0.001 |
| | Piecewise[c] | 4537.7 | -2112.2 | 71.80 | 18 | < 0.001 |
| MoCA | Linear | 4513.4 | -2172.0 | | | |
| | Quadratic | 4538.1 | -2164.0 | 15.94 | 6 | 0.014 |
| | Piecewise | 4603.0 | -2155.8 | 32.44 | 18 | 0.019 |
| Memory | Linear | 1962.9 | -897.0 | | | |
| | Quadratic | 1979.0 | -884.7 | 24.48 | 6 | < 0.001 |
| | Piecewise | 2048.7 | -879.0 | 35.87 | 18 | 0.007 |
| Executive Function | Linear | 1600.1 | -655.4 | | | |
| | Quadratic | 1650.4 | -640.3 | 30.39 | 12 | 0.002 |
| | Piecewise | 1776.8 | -622.8 | 65.36 | 36 | 0.002 |
| Visuospatial Function | Linear | 2785.0 | -1246.7 | | | |
| | Quadratic | 2842.4 | -1234.8 | 23.95 | 12 | 0.021 |
| | Piecewise | 2981.0 | -1222.7 | 48.10 | 36 | 0.086 |
| Processing Speed | Linear | 1817.1 | -766.2 | | | |
| | Quadratic | 1869.1 | -752.5 | 27.48 | 12 | 0.007 |
| | Piecewise | 2000.1 | -738.6 | 55.29 | 36 | 0.021 |
| Language | Linear | 2248.0 | -1038.6 | | | |
| | Quadratic | 2269.9 | -1029.0 | 19.11 | 6 | 0.004 |
| | Piecewise | 2338.1 | -1022.1 | 32.91 | 18 | 0.017 |

**Note.** Abbreviations: BIC = Bayesian Information Criterion; CDR-SB = Clinical Dementia Rating-Sum of Boxes; df = degrees of freedom; MoCA = Montreal Cognitive Assessment. Better model fit was indicated by lower BIC and higher log-likelihood values. $\chi^2$, df, and *p*-values were derived from likelihood ratio tests comparing models with non-linear and linear baseline pTau217 parameterizations for the same clinical outcome. Significant tests suggested the more complex model was a better fit.

[a]Original model including only the linear baseline pTau217 term.

[b]Model including linear and quadratic baseline pTau217 terms.

[c]Model including linear and piecewise baseline pTau217 terms.

**eTable 6.** Simple slope estimates from linear mixed-effects models with and without a quadratic baseline pTau217 term in LBD

| Clinical Outcome | Simple Slope Effect in LBD | Model including quadratic baseline pTau217 | | | | | Model including linear baseline pTau217 only | | | | |
|---|---|---|---|---|---|---|---|---|---|---|---|
| | | Est. | SE | 95% CI | | *p*-value | Est | SE | 95% CI | | *p*-value |
| CDR-SB | Linear Baseline pTau217 | 0.38 | 0.09 | 0.21 | 0.55 | < 0.001 | 0.30 | 0.08 | 0.14 | 0.45 | < 0.001 |
| | pTau217 Slope | 0.20 | 0.08 | 0.05 | 0.35 | 0.025 | 0.24 | 0.07 | 0.10 | 0.38 | 0.003 |
| MoCA | Linear Baseline pTau217 | -0.47 | 0.12 | -0.71 | -0.23 | < 0.001 | -0.35 | 0.11 | -0.57 | -0.13 | 0.002 |
| | pTau217 Slope | -0.01 | 0.11 | -0.23 | 0.22 | 0.942 | -0.06 | 0.12 | -0.30 | 0.17 | 0.595 |
| Memory | Linear Baseline pTau217 | -0.10 | 0.03 | -0.15 | -0.05 | 0.001 | -0.06 | 0.02 | -0.11 | -0.02 | 0.015 |
| | pTau217 Slope | -0.02 | 0.02 | -0.07 | 0.03 | 0.946 | -0.03 | 0.03 | -0.08 | 0.02 | 0.423 |
| Executive | Linear Baseline pTau217 | -2.26 | 0.46 | -3.17 | -1.35 | < 0.001 | -1.47 | 0.39 | -2.24 | -0.70 | 0.001 |
| | pTau217 Slope | -0.66 | 0.37 | -1.39 | 0.06 | 0.110 | -0.78 | 0.40 | -1.57 | 0.02 | 0.082 |
| Visuospatial | Linear Baseline pTau217 | -4.19 | 1.03 | -6.22 | -2.17 | < 0.001 | -2.15 | 0.83 | -3.78 | -0.53 | 0.028 |
| | pTau217 Slope | -0.05 | 0.82 | -1.65 | 1.55 | 0.95 | -0.26 | 0.86 | -1.95 | 1.43 | 0.762 |
| Processing | Linear Baseline pTau217 | -1.92 | 0.67 | -3.23 | -0.61 | 0.012 | -1.15 | 0.54 | -2.20 | -0.10 | 0.097 |
| | pTau217 Slope | -0.18 | 0.54 | -1.23 | 0.88 | 0.743 | -0.29 | 0.57 | -1.41 | 0.83 | 0.615 |
| Language | Linear Baseline pTau217 | -0.01 | 0.02 | -0.06 | 0.04 | 0.597 | -0.01 | 0.02 | -0.05 | 0.03 | 0.658 |
| | pTau217 Slope | -0.03 | 0.02 | -0.07 | 0.02 | 0.643 | -0.03 | 0.02 | -0.07 | 0.01 | 0.355 |

**Note.** Abbreviations: CDR-SB = Clinical Dementia Rating-Sum of Boxes; Executive = Executive function; Est. = Simple slope estimate; LBD = Lewy body disease; MoCA = Montreal Cognitive Assessment; Processing = Processing speed; SE = Standard error; Visuospatial = Visuospatial function. Estimates represent simple slope effects of either baseline pTau217 or pTau217 slope on longitudinal change in clinical outcome in the LBD group. Models including the quadratic baseline pTau217 term also retained the linear baseline pTau217 and pTau217 slope variables.

**eTable 7.** Unadjusted Kaplan-Meier estimates for time to conversion to MCI or dementia stratified by baseline pTau217 status and clinical diagnosis

| Conversion Event | Survival Group | *N* | Events *n* (%)[a] | Time to Conversion[b] | 5-Year Survival (%) |
|---|---|---|---|---|---|
| MCI or Dementia[c] (*n* = 371) | HC pTau217- | 142 | 9 (6.33) | - | 92.50 |
| | HC pTau217+ | 78 | 16 (20.51) | 7.99 | 80.91 |
| | AD pTau217- | 20 | 6 (30.00) | 6.99 | 54.97 |
| | AD pTau217+ | 35 | 14 (40.00) | 5.25 | 52.20 |
| | LBD pTau217- | 73 | 23 (31.50) | 7.26 | 76.30 |
| | LBD pTau217+ | 23 | 12 (52.17) | 2.96 | 36.68 |
| MCI or Dementia (*n* = 275) | HC pTau217- | 137 | 6 (4.37) | - | 95.3 |
| | HC pTau217+ | 77 | 16 (20.78) | 7.99 | 85.6 |
| | PDNC pTau217- | 51 | 18 (35.29) | 7.15 | 74.1 |
| | PDNC pTau217+ | 10 | 6 (60.00) | 2.93 | 41.4 |
| Dementia[d] (*n* = 36) | MCI-PD pTau217- | 17 | 1 (5.88) | 7.96 | 100.0[d] |
| | MCI-PD pTau217+ | 6 | 2 (33.33) | 4.89 | 75.0[d] |
| | MCI-LB pTau217- | 6 | 5 (83.33) | 4.23 | 66.7[d] |
| | MCI-LB pTau217+ | 7 | 4 (57.14) | 2.97 | 35.7[d] |

**Note.** Abbreviations: AD = Alzheimer’s disease spectrum; HC = Healthy control; LBD = Lewy body disease spectrum; MCI = Mild cognitive impairment; MCI-LB = MCI due to Lewy body disease (prodromal dementia with Lewy bodies); MCI-PD = MCI due to Parkinson’s disease; PDNC = Parkinson’s disease with normal cognition; pTau217+ = Baseline plasma pTau217 > 0.54pg/mL (abnormal); pTau217- = Baseline plasma pTau217 ≤ 0.54pg/mL (normal). Five-year survival reflects proportion remaining free or cognitive conversion after 5 years.
[a]Percentage based on number of participants in survival group.
[b]Median time to conversion in years, not estimated when fewer than 50% experienced the event (-).
[c]Conversion defined as progression from cognitively unimpaired to MCI or MCI to dementia in HC and LBD and progression from MCI to dementia in AD.
[d]3-year survival probability for conversion to dementia due to sample size.

**eTable 8.** Cox models for diagnostic progression to MCI or dementia stratified by baseline cognitive status ($n$ = 371)

| Conversion Event | Model Predictor | Hazard Ratio | 95% CI | | $p$-value |
|---|---|---|---|---|---|
| MCI or Dementia | HC pTau217- | 1.00 | - | - | - |
| | HC pTau217+ | 3.26 | 1.44 | 7.40 | 0.005 |
| | AD pTau217- | 9.73 | 2.71 | 34.97 | 0.001 |
| | AD pTau217+ | 14.65 | 4.87 | 43.53 | < 0.001 |
| | LBD pTau217- | 5.77 | 2.57 | 12.93 | < 0.001 |
| | LBD pTau217+ | 21.96 | 8.19 | 58.88 | < 0.001 |
| | Age | 1.03 | 0.99 | 1.06 | 0.095 |
| | Sex | 1.01 | 0.62 | 1.64 | 0.966 |

**Note.** Abbreviations: AD = Alzheimer's disease spectrum; HC = Healthy control; LBD = Lewy body disease spectrum; MCI = Mild cognitive impairment; pTau217+ = Baseline plasma pTau217 ≥ 0.54pg/mL (abnormal); pTau217- = Baseline plasma pTau217 < 0.54pg/mL (normal). Hazard ratios were derived from Cox models stratified by baseline cognitive status (cognitively unimpaired versus impaired), thereby allowing baseline hazards to differ across strata and accounting for differences in conversion risk between individuals with no cognitive impairment and MCI at baseline in LBD. Hazard ratios >1 indicate greater hazard of diagnostic progression relative to the reference group. In LBD and HC, conversion was defined as progression from cognitively unimpaired to MCI or MCI to dementia. In AD, conversion was defined as progression from MCI to dementia. Results are reported relative to HC pTau217-.

#### 5. eFigures

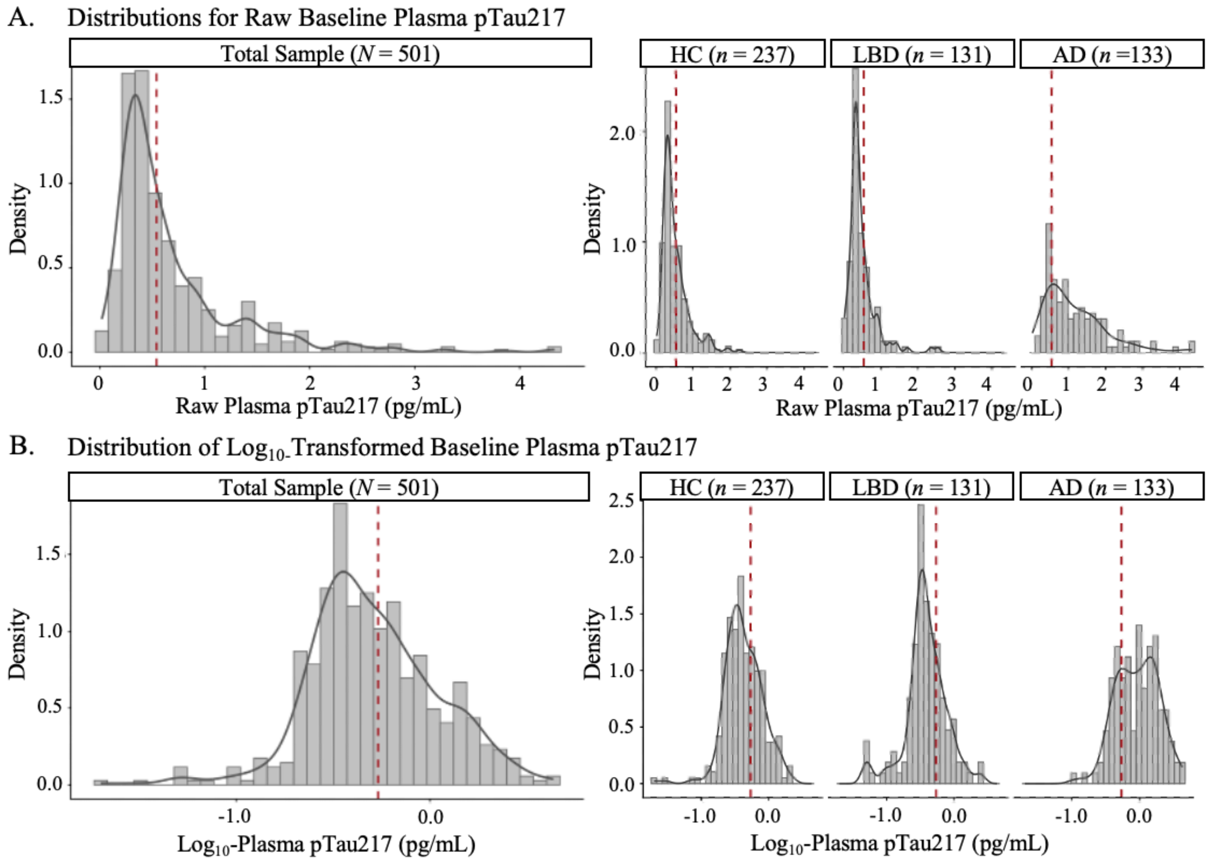


**eFigure 1.** Distributions for raw and $\log_{10}$-transformed baseline plasma pTau217.

Distributions for raw and $\log_{10}$-transformed baseline plasma pTau217 are shown in panels (A.) and (B.), respectively, for the total sample (*N* = 501) on the left and stratified by clinical diagnosis group (HC, *n* = 237; LBD, *n* = 131; AD, *n* = 133) on the right. Density curves are overlaid in solid dark-grey lines and the LBD-specific cut-point of 0.54pg/mL is shown as a vertical dashed red line. AD = Alzheimer's disease spectrum; HC = Healthy control; LBD = Lewy body disease spectrum.

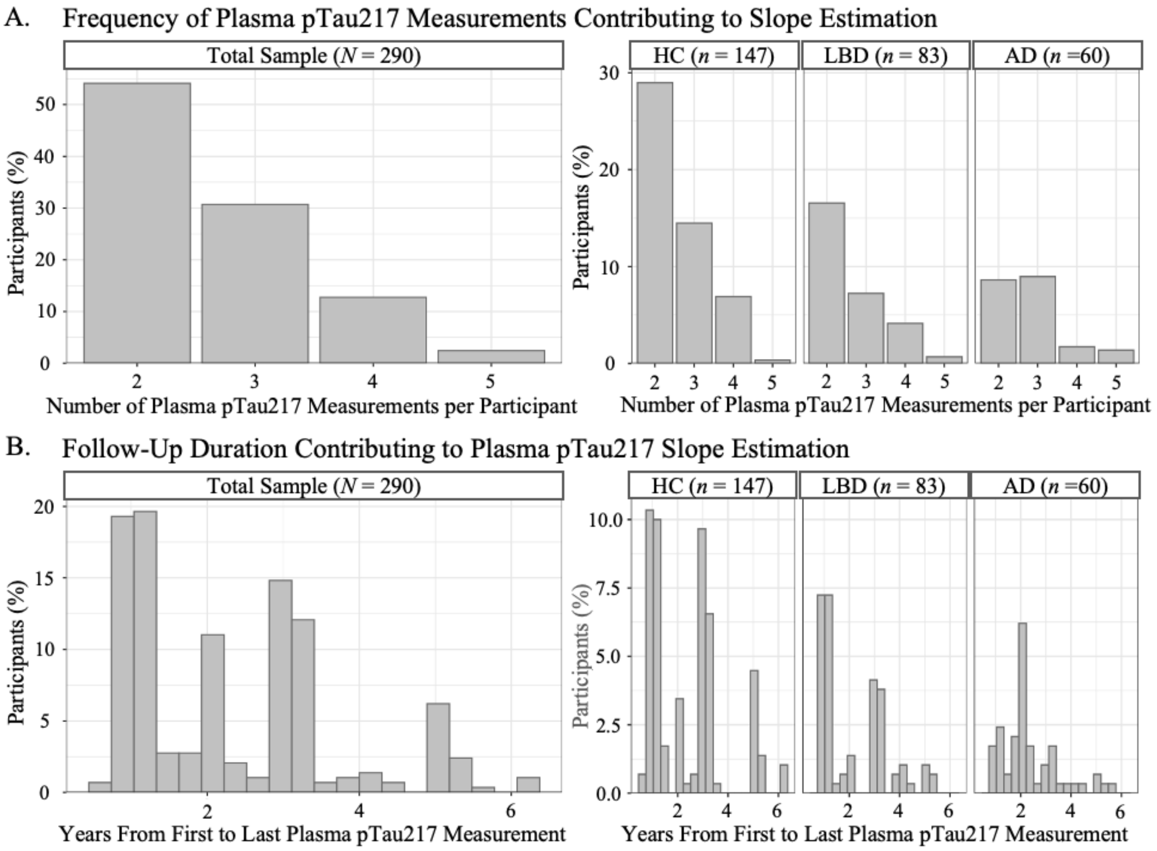


**eFigure 2.** Plasma pTau217 measurement frequencies and follow-up duration contributing to slope estimation.

Panel (A.) shows distributions for total number of plasma pTau217 measurements per participant in the total sample (i.e., participants with ≥ 2 plasma measurements; *N* = 290) on the left and stratified by clinical diagnosis group on the right (HC, *n* = 147; LBD, *n* = 83; AD, *n* = 60). Panel (B.) shows distributions for total follow-up duration (i.e., years between first and last plasma pTau217 measurement) per participant in the total sample on the left and stratified by clinical diagnosis group on the right. The y-axis in both panels represents percentage of participants in the total sample or within each clinical diagnosis group. AD = Alzheimer's disease spectrum; HC = Healthy control; LBD = Lewy body disease spectrum.

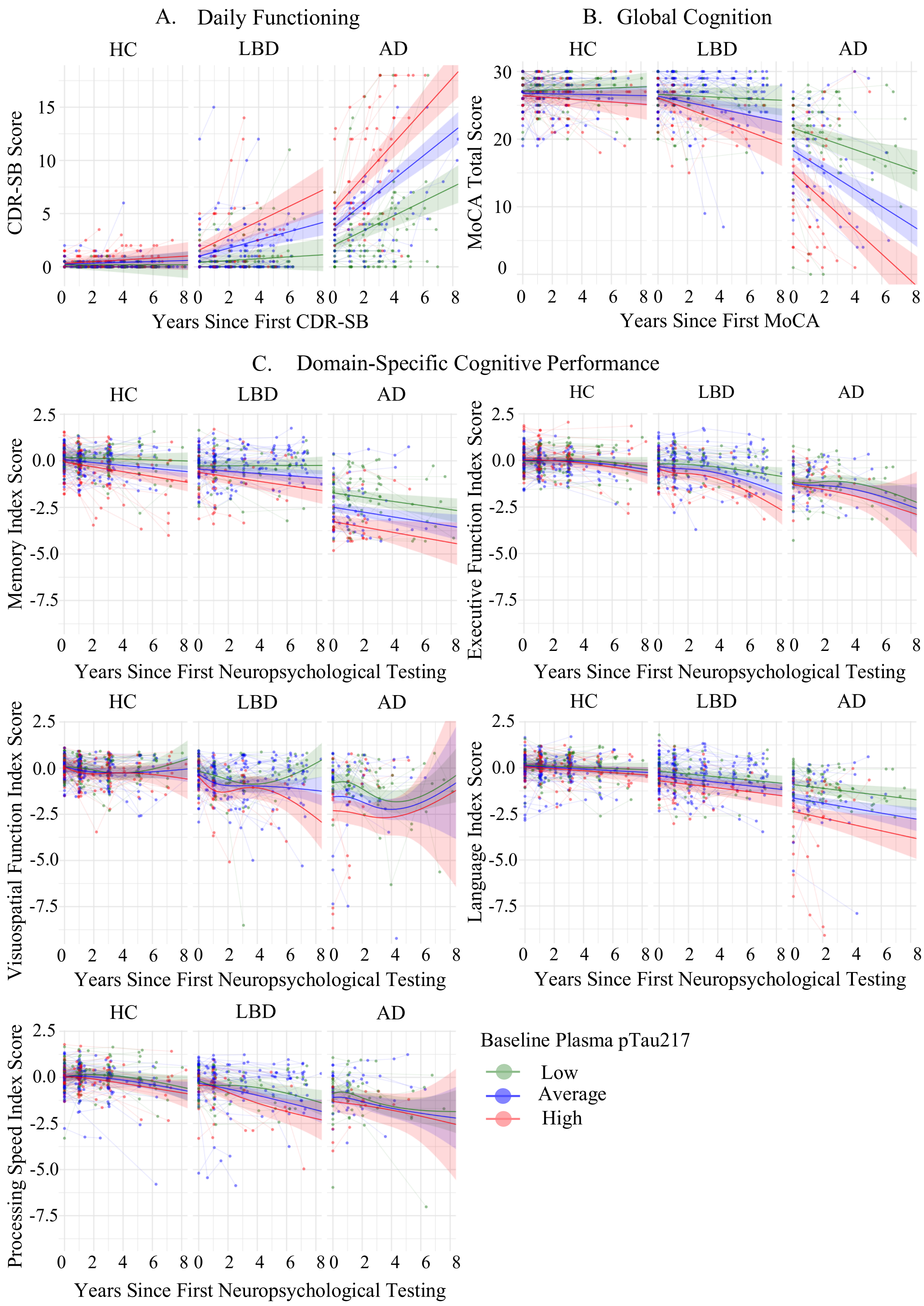
A. Daily Functioning
B. Global Cognition
HC
LBD
AD
CDR-SB Score
Years Since First CDR-SB
MoCA Total Score
Years Since First MoCA
C. Domain-Specific Cognitive Performance
Memory Index Score
Executive Function Index Score
Visuospatial Function Index Score
Language Index Score
Processing Speed Index Score
Years Since First Neuropsychological Testing
Baseline Plasma pTau217
Low
Average
High

**eFigure 3.** Effect of baseline plasma pTau217 on longitudinal clinical outcome trajectories.

Observed participant (thin lines) and model-estimated (bold lines with associated 95% confidence interval shading) trajectories are shown for (A.) CDR-SB score, (B.) MoCA total score, and (C.) five cognitive index scores for each clinical diagnosis group (HC, LBD, AD) over 2-8 years. Points represent individual clinical outcome observations. For visualization purposes only, trajectories were grouped by baseline plasma pTau217 level relative to the total sample (*n* = 290) mean where low = -1 standard deviation (SD; green), average = 0 SD (blue), and high = +1 SD (red). pTau217 slope was held at 0 SD to isolate baseline pTau217 effects. AD = Alzheimer's disease spectrum; CDR-SB = Clinical Dementia Rating-Sum of Boxes; HC = Healthy control; LBD = Lewy body disease spectrum; MoCA = Montreal Cognitive Assessment.

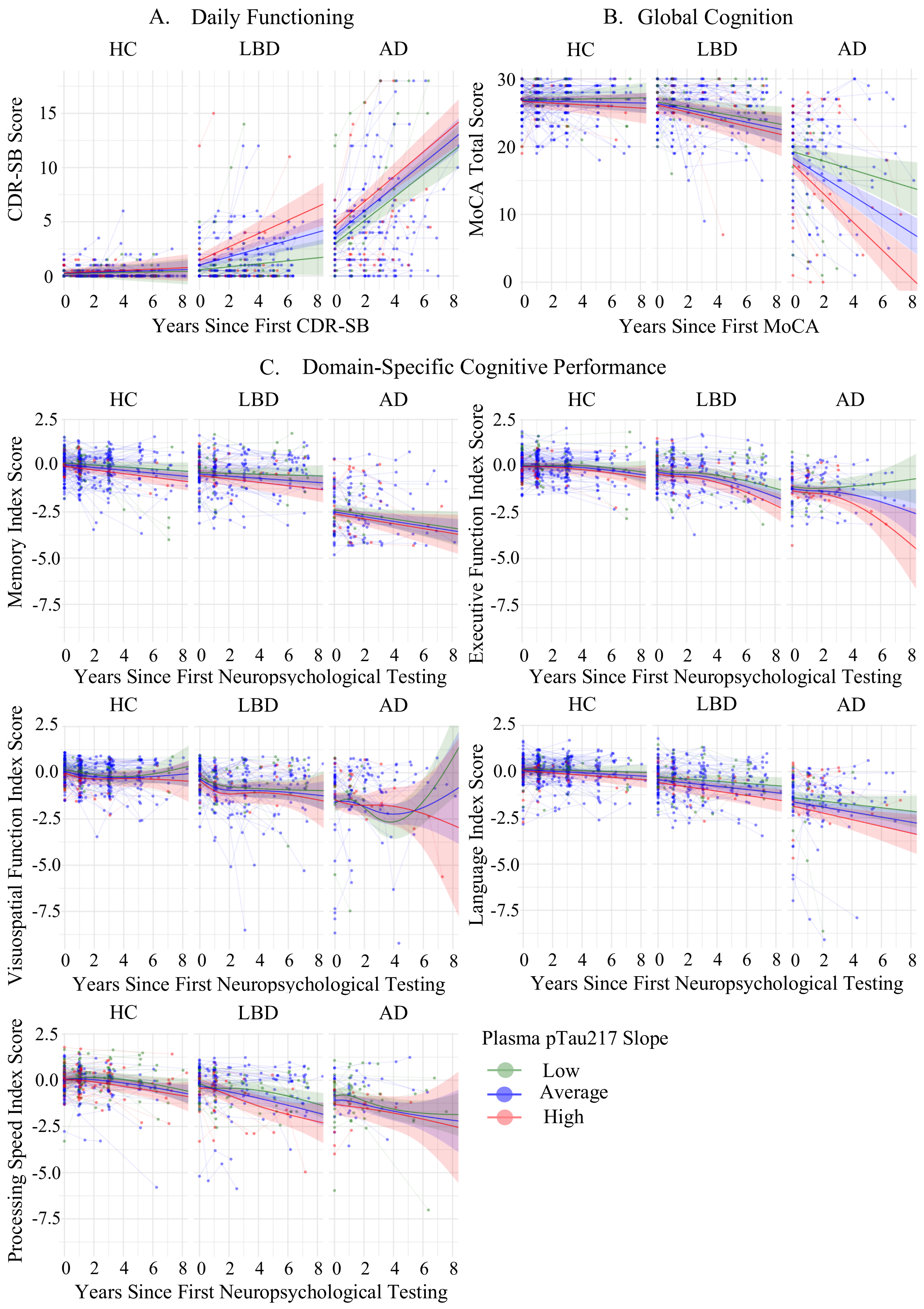
A. Daily Functioning
HC
LBD
AD
CDR-SB Score
Years Since First CDR-SB
B. Global Cognition
MoCA Total Score
Years Since First MoCA
C. Domain-Specific Cognitive Performance
Memory Index Score
Executive Function Index Score
Visuospatial Function Index Score
Language Index Score
Processing Speed Index Score
Years Since First Neuropsychological Testing
Plasma pTau217 Slope
Low
Average
High

**eFigure 4.** Effect of plasma pTau217 slope on longitudinal clinical outcome trajectories.

Observed participant (thin lines) and model-estimated (bold lines with associated 95% confidence interval shading) trajectories are shown for (A.) CDR-SB score, (B.) MoCA total score, and (C.) five cognitive index scores for each clinical diagnosis group (HC, LBD, AD) over 2-8 years. Points represent individual clinical outcome observations. For visualization purposes only, trajectories were grouped by plasma pTau217 slope relative to the total sample ($n$ = 290) mean where low = -1 standard deviation (SD; green), average = 0 SD (blue), and high = +1 SD (red). Baseline pTau217 was held at 0 SD to isolate pTau217 slope effects. AD = Alzheimer's disease spectrum; CDR-SB = Clinical Dementia Rating-Sum of Boxes; HC = Healthy control; LBD = Lewy body disease spectrum; MoCA = Montreal Cognitive Assessment.
